# Exposure accumulation drives age-dependent disease architectures and polygenic risk scores

**DOI:** 10.1101/2025.08.29.25334745

**Authors:** Xilin Jiang, Alkes L. Price, John Danesh, Gil McVean, Michael Inouye, Sarah Urbut, Arun Durvasula

**Affiliations:** British Heart Foundation Cardiovascular Epidemiology Unit, Department of Public Health and Primary Care, University of Cambridge, Cambridge UK; Victor Phillip Dahdaleh Heart and Lung Research Institute University of Cambridge, Cambridge UK; Department of Epidemiology, Harvard T.H. Chan School of Public Health Boston MA USA; Broad Institute, Cambridge, MA USA; Cambridge Centre for AI in Medicine, University of Cambridge, Cambridge UK; Department of Biostatistics, Harvard T.H. Chan School of Public Health Boston MA USA; British Heart Foundation Centre of Research Excellence, University of Cambridge, Cambridge, UK; National Institute for Health and Care Research Blood and Transplant Research Unit in Donor Health and Behaviour, University of Cambridge, Cambridge, UK; Health Data Research UK Cambridge, Wellcome Genome Campus and University of Cambridge, Cambridge, UK; Department of Human Genetics, Wellcome Sanger Institute, Hinxton, UK; Ellison Institute of Technology, Oxford, UK; Cambridge Baker Systems Genomics Initiative, Department of Public Health and Primary Care, University of Cambridge, Cambridge, UK; Cambridge Baker Systems Genomics Initiative, Baker Heart and Diabetes Institute, Melbourne, Victoria, Australia; Cardiology Division, Massachusetts General Hospital, Boston, MA USA; Division of Epidemiology, Department of Population and Public Health Sciences, Keck School of Medicine, University of Southern California, Los Angeles, CA; Center for Genetic Epidemiology, Department of Population and Public Health Sciences, Keck School of Medicine, University of Southern California, Los Angeles, CA; Department of Quantitative and Computational Biology, University of Southern California, Los Angeles, California, USA

## Abstract

Our understanding of the dependence of the genetic and environmental architecture of common diseases on age is incomplete. Here, we use longitudinal data to separate age-dependent genetic and environmental variance across complex traits and diseases. When applying our approach to 16 UK Biobank quantitative traits (average *N=*180K), we found environmental variance accumulates with age, leading to decreasing heritability with age, which follows an exposure accumulation model that is distinct from gene-age interaction model. Heritability decreases with age for 5 traits including systolic blood pressure and lung function (FEV1/FVC), with an average change of −17.8% with age per 10-years. We demonstrated that majority of the decreasing heritability comes from exposure accumulation, instead of gene-age interaction, using longitudinal phenotypic correlations. For diseases, environmental variance in disease liability also accumulates with age for 5 of 9 diseases, with an average decrease in heritability of −18.1% per 10 years. A liability-threshold model with exposure accumulation explains 86% of decreasing PRS prediction R^2^ with age in 9 UK Biobank complex diseases. Finally, we show that both genetic and non-genetic predictors have decreasing prediction accuracy with age in 61 predictor-disease pairs. Taken together, our results demonstrate that the age-dependent liability-threshold model with exposure accumulation is a general disease model, suggesting ascertaining younger cohorts is more powerful for training both genetic and non-genetic risk prediction models.

## Introduction

The extent to which genetics and environmental exposures interact with age when contributing to phenotypic variation in a population is unclear^1,2^. Previous studies have found evidence that environmental exposures can account for more phenotypic variation in older individuals, potentially due to the accumulation of exposures as people age^3–5^. However, measuring the relevant environmental exposures throughout an individual’s lifetime remains a significant challenge. On the other hand, gene-age interaction, which can arise from multiple sources^6^, including proportional amplification of genetic variance^7^ and different age profiles of genetic effects across genotypes^8^, could drive age-dependent phenotypic variation. Jointly studying the age profiles of genetic and non-genetic components of complex traits can elucidate the architectures of age-dependent complex traits.

Recent work has shown age-varying prediction accuracy for polygenic risk scores (PRS), but whether this observation is driven by gene-age interactions or other mechanisms is unknown^8–14^. Significant challenges remain in studying age-dependent disease architectures. First, disease liability is not observed and performing gene-age interaction tests on binary disease observations may not be informative of interactions on the liability scale^15^. Second, although methods exist for reducing bias in modeling time-to-event data due to censoring, these methods often have assumptions that are not met in empirical data^8^.

Here, we estimated the age-dependent profiles of genetic and environment components of complex traits and disease liability. We show that the impact of environmental exposures accumulates with age for a wide range of complex traits and diseases, which causes a decrease in heritability with age. We show that exposure accumulation, under a liability-threshold model^16,17^, explains decreasing prediction accuracy of PRS prediction accuracy for 9 diseases in UK Biobank. Finally, we show by simulation and analytical derivation that a liability-threshold model predicts that both genetic and environmental factors will have decreased prediction accuracy for older cases, which we validate in empirical data using 61 predictor-disease pairs.

## Results

### Overview of methods

We study age-dependent trait architecture under an exposure accumulation (EA) model where liability is linear in genetic, environment, and age quintiles^18^, with environmental variance accumulating with age.

The EA model can be formalized under the following:

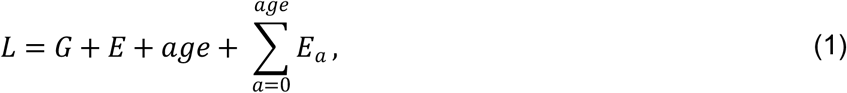

where *L* denotes liability, *G* is the genetic component of liability, *E* is the environmental component, *age* is the quintile of age, and *E*_*a*_ is the contribution of age to liability. 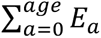 is the term arising from the EA model. We study architecture across age quintiles to relax statistical assumptions on the relationship between age and *L*. We evaluate evidence for 4 additional models that can cause an age-dependent architecture: proportional amplification^7,15^ (ref), age-dependent G, exposure accumulation with decay, and non-proportional amplification (**Supplementary Table 1**).

We evaluate evidence of the EA model by estimating age-varying heritability 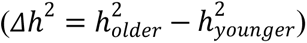, which is negative under the EA model. We estimate *Δh*^2^ by estimating the heritability for each age quintile with BOLT-REML^19^ and fitting a model where heritability changes linearly with age. We estimate heritability with age and sex as covariates to account for within age quintile variation due to age and sex. We test for non-zero *Δh*^2^ with a likelihood ratio test between the model with age term and a null model where heritability is constant with age. We also evaluate the environmental correlation between age quintiles (*ρ*(*E*_*younger*_, *E*_*older*_)), which is expected to be less than 1.0 under the EA model. We use phenotypic correlations (*ρ*_*l*_), which is an upper bound of *ρ*(*E*_*younger*_, *E*_*older*_) under the EA model, to evaluate evidence of *ρ*(*E*_*younger*_, *E*_*older*_) < 1. We account for error-in-measurement of *ρ*_*l*_ by computing the denoised phenotypic correlation 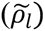 for individuals with three phenotype measurements at different age points. We estimated 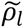 as the ratio of first-third visit *ρ*_*l*_ and first-second visit *ρ*_*l*_. The ratio cancels the impact of measurement error. We divide 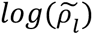 by the average age difference between the second and third visits to estimate 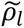 between individuals with one year of age difference (see **Methods**).

We estimated *Δh*^2^ and 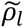 for 16 quantitative traits across 180K individuals in the UK Biobank (**Supplementary Table 2**); the sample size reflects a restriction to individuals of British ancestry with diagnostic records from both primary care data and hospital inpatient data, which we imposed to mitigate the impact of missing diagnoses. The quantitative traits in this study were selected following ref. ^20^ (**Methods**). We use *cross-sectional design* to estimate *Δh*^2^, where we compare individuals with different ages at the baseline visit. We evaluate the impact of recruitment bias that can arise from a cross-sectional design in a secondary analysis. We use *longitudinal measurement* to estimate 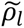, where we compare multiple measurements at different age points of the same individual.

We estimate *Δh*^2^ for the estimated disease liability of 9 diseases (**Supplementary Table 3**). We use 36 quantitative traits (**Supplementary Table 4**) to estimate disease liability at baseline age by predicting binary disease phenotypes from the quantitative traits. We do not estimate 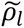 for estimated disease liability because we do not have longitudinal measurements for all quantitative traits that were used to estimate disease liability.

We study the impact of age-dependent disease architecture on disease prediction using simulations. We extract age-at-diagnosis for each patient using the first diagnosis in either primary care data or hospital inpatient data to evaluate age-dependent disease prediction accuracy in empirical data. We construct PRS and non-genetic predictors using five-fold cross validation without age stratification. We have publicly released summary data of age-specific heritability, phenotypic correlation, and genetic correlation estimates (see **Data Availability**). We have also released software to compute age-dependent heritability and prediction accuracy (and **Code Availability**).

### Age-dependent genetic architecture of quantitative traits

We estimated *Δh*^2^ w.r.t. age of 16 quantitative traits (**Supplementary Table 2**) to evaluate the evidence supporting the EA model. First, we found 5 of the 16 traits have significantly decreasing heritability with age (*p*<0.05/9, FEV1/FVC, LDL cholesterol, total cholesterol, diastolic blood pressure, and systolic blood pressure) with an average 10-year decrease of 17.8% in *h*^2^ (**Figure 1A**, **Supplementary Table 5, Supplementary Figure 1**). Second, we computed genetic and environmental variance of each age quintile using heritability and phenotypic variance estimates (**Figure 1B, Supplementary Figure 2, Supplementary Figure 3, Supplementary Table 6, Supplementary Table 7**). We estimated phenotypic variance for males and females separately to account for sex differences^7,15^, which provides estimates for 32 trait-sex pairs. We found *Δvar*(*E*) is significantly larger than *Δvar*(*G*) for 10 trait-sex pairs (red dots in **Figure 1B**), supporting the EA model. We found *var*(*E*) and *var*(*G*) vary proportionally with age in traits that do not have significant age-varying heritability (grey dots in **Figure 1B**), supporting a proportional amplification model that has been observed in GxE studies^7,15^. The EA model will move traits above the diagonal line in **Figure 1B**, while the proportional amplification model will move traits along the diagonal line. Taken together, we conclude that FEV1/FVC, LDL cholesterol, total cholesterol, systolic blood pressure, and diastolic blood pressure show evidence for the EA model.

**Figure 1.**
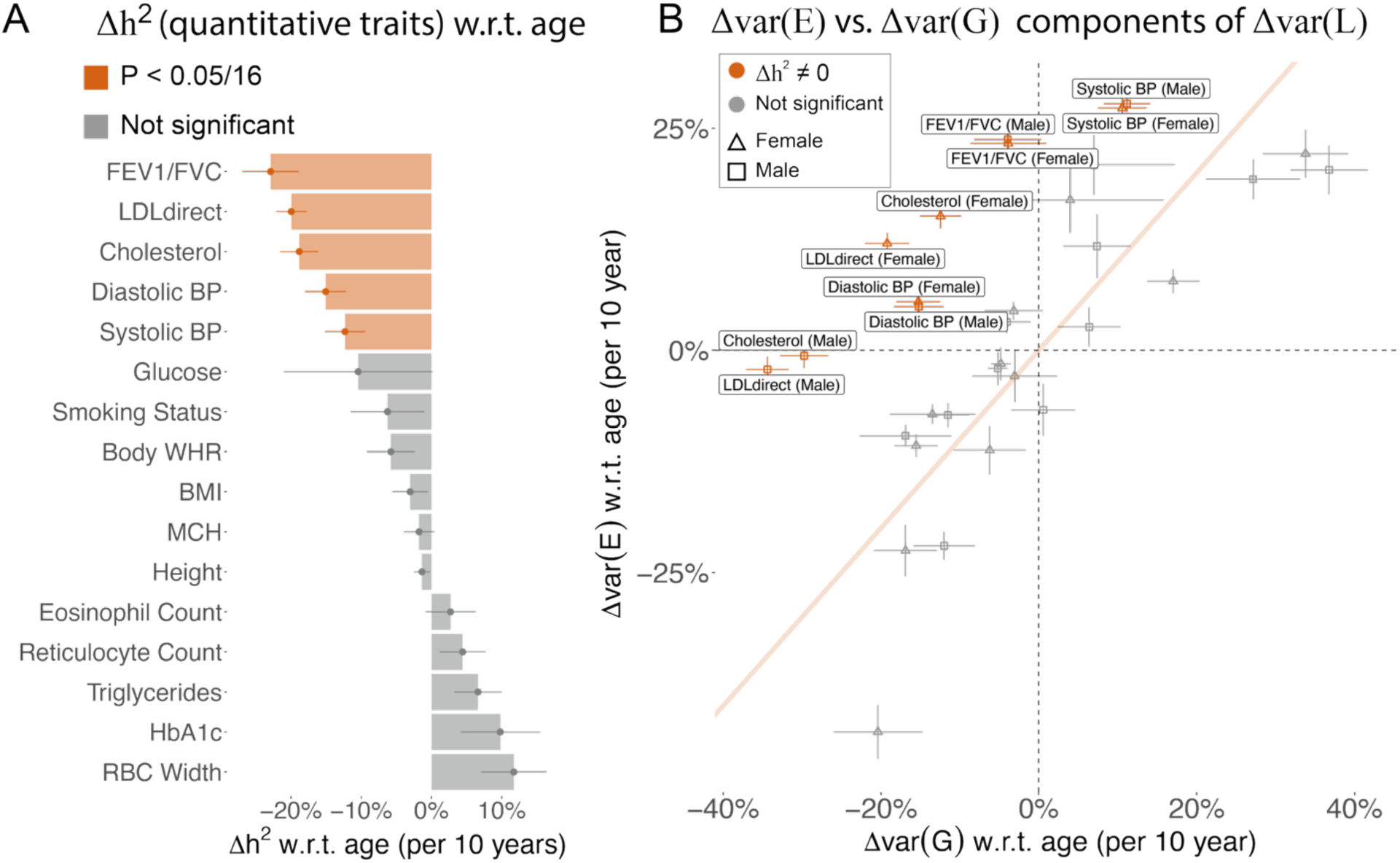
Age-dependent variance of quantitative traits. (A) Heritability changes per 10-years for 16 quantitative traits. Red bars show significant heritability changes across age; error bars show standard error via Monte-Carlo sampling. (B) 10-year change for genetic and environment variance for the 16 quantitative traits. Traits with significant age-dependent heritability are highlighted in red. Traits in grey show evidence of proportional amplification of genetic and environmental variance. Error bars are standard error via Monte-Carlo sampling. Numerical results are provided in **Supplementary Table 5 and Supplementary Table 7**. We note that no estimates are significantly outside the plausible range and that allowing estimates outside of the plausible range is necessary to ensure unbiasedness of estimates.

Next, we estimated genetic and phenotypic correlations between age groups to evaluate additional evidence in support of the EA model. *ρ*(*G*_*younger*_, *G*_*older*_) is expected to equal 1.0 and *ρ*(*E*_*younger*_, *E*_*older*_) is expected to be less than 1.0 under the EA model. We found the genetic correlation, *ρ*(*G*_*younger*_, *G*_*older*_), was close to 1.0 for most traits, with exceptions of HbA1c (*p*=1e-4) and glucose (*p*=3e-4) (**Figure 2A, Supplementary Table 8**). A genetic correlation less than 1.0 supports a model where genetic components have different age profiles (**Supplementary Table 1**), which is consistent with our previous work that showed that genetic variants have distinct age profiles in conveying common disease risk^4^. We computed the denoised phenotypic correlation 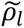 to evaluate the environmental correlation across age (*ρ*(*E*_*younger*_, *E*_*older*_)) (**Methods**). For the 11 traits that have longitudinal data for 3 visits, 7 have 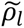 significantly less than 1.0 (**Figure 2B, Supplementary Table 9**), supporting the EA model. 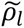 has high correlation with *Δh*^2^ w.r.t. age (*ρ*=0.61 across 11 traits, **Figure 2C**), which is consistent with the EA model. Specifically, the 3 traits (Systolic BP, Diastolic BP, and FEV1/FVC) that show evidence supporting the EA model in **Figure 1** have the largest difference between 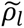 and 1. Our analysis of the genetic and environmental correlation between young and old individuals provides orthogonal evidence in support of EA for systolic blood pressure, diastolic blood pressure, and FEV1/FVC.

**Figure 2.**
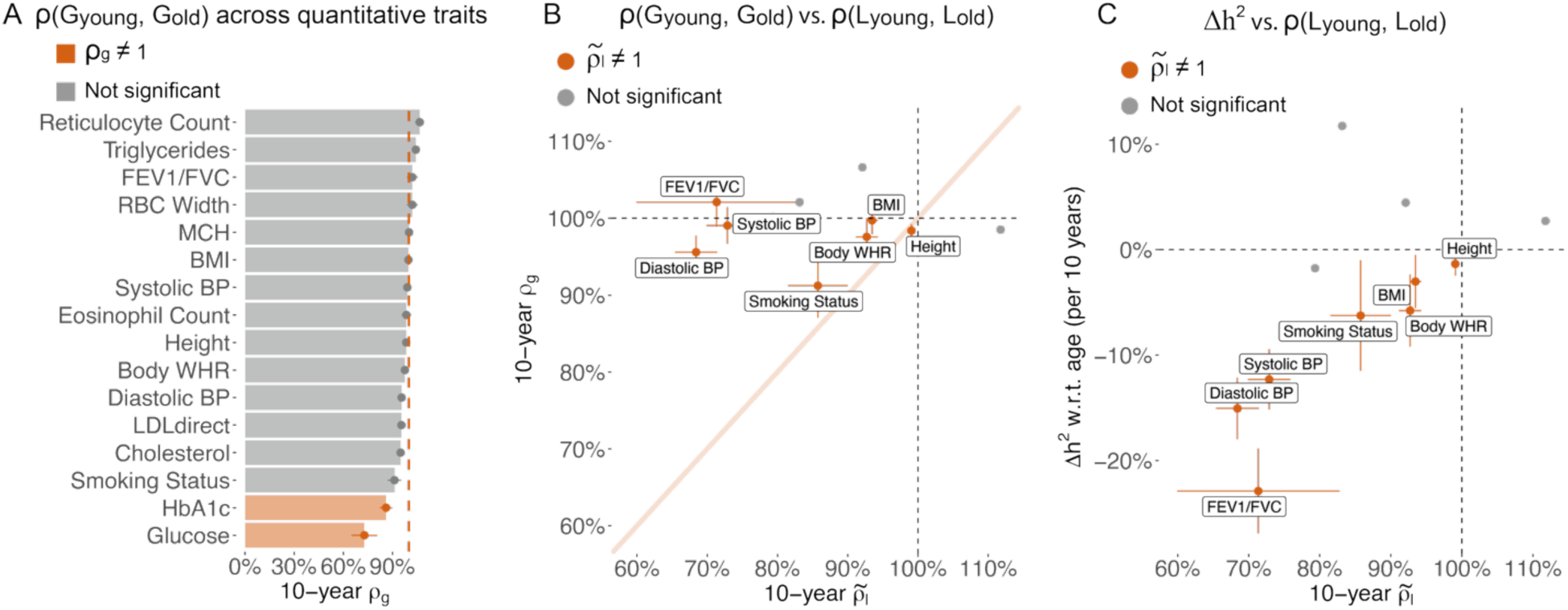
Genetic liability is more correlated across young and older age groups than environmental liability. (A) 10-year genetic correlation between younger and older age bins for 16 quantitative traits. We report the genetic correlation per 10-year age difference. (B) Scatter plot of 10-year genetic correlation versus denoised 10-year phenotypic correlation for the 11 quantitative traits that have three measurements per individual. We found no genetic correlation or phenotypic correlation that are significantly higher than 1 at nominal *p*-value = 0.05. (C) Scatter plot of heritability changes per 10-years versus denoised 10-year phenotypic correlation for the 11 quantitative traits that have three measurements per individual. Traits with denoised 10-year phenotypic correlation significantly different from 1 are shown in red and labeled in panel B and C; non-significant traits are shown in grey. Error bars for phenotypic correlation are standard error via bootstrapping over individuals; error bars for genetic correlation are standard error after meta-analysis across 4 age-bin pairs. Numerical results are provided in **Supplementary Table 8-9**. We note that no estimates are significantly outside the plausible range and that allowing estimates outside of the plausible range is necessary to ensure unbiasedness of estimates.

We performed six secondary analyses to test the robustness of our results. First, we tested whether age-dependent recruitment bias of individuals with different health status could explain decreasing heritability estimates. We created two age-matched samples (mean age: 55.9 vs. 56.6) with high and low numbers of prevalent disease diagnoses (mean diagnoses: 6.8 vs 34.6) and estimated the heritability of 16 quantitative traits in the two groups. We observed minimal heritability differences between the two groups, which suggests that biased recruitment of healthier older individuals in the UK Biobank^21^ does not contribute to the decreasing heritability estimates with age (**Supplementary Figure 4**). Second, we evaluated the amplification models^7^ by estimating changes in genetic variance w.r.t. age for 32 trait-sex pairs (**Supplementary Figure 2, Supplementary Table 7**). We found 25 traits with age-varying genetic variance, align with previous GxSex and GxE studies that report amplifications^7,15^. Third, we tested an alternative model where exposure accumulation decays with age by identifying traits with *Δh*^2^ close to zero but a denoised phenotypic correlation less than 1.0 across age bins. We found evidence support this model for BMI and body WHR among the traits that we can estimate 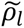 (i.e. those with multiple measurements) (**Supplementary Table 9**). Fourth, we tested a non-proportional amplification model by identifying traits with both genetic correlation and denoised phenotypic correlation close to 1 but age-varying heritability (**Supplementary Table 9**). We found no such traits among the traits that we can estimate 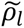. Fifth, we estimated genetic correlation between the top and bottom age quantiles (median age difference = 22.0) without meta-analyzing across age bins and found smaller *ρ*(*G*_*young*_, *G*_*old*_) compared with the meta-analyzed results with larger standard errors (**Supplementary Figure 5**). Sixth, we tested whether using age in years instead of age quintiles can explain more variance in quantitative traits. We found age quintiles explains similar *R*^2^ as age in years (**Supplementary Figure 6A**), suggesting age quintiles capture the majority of age-related variation while allowing us to use standard methods to compute heritability and genetic correlations. We found adding an additional age^2^ covariate to the age in years explains more quantitative trait variance (mean *R*^2^= 0.024 across 16 traits) than the model with only linear age in years (mean *R*^2^= 0.022 across 16 traits).

Taken together, these results support exposure accumulation as the primary driver of age-dependent architecture in quantitative traits.

### Age-dependent genetic architecture of estimated disease liability

We next investigated whether the liability of 9 diseases (**Supplementary Table 3**) is consistent with the EA model. We estimated disease liability by applying LASSO^22^ to 36 quantitative traits (**Supplementary Table 4**) measured at baseline and predicting disease status in the entire population (i.e. across all age bins). We use the value predicted by LASSO as estimated liability to approximate disease liability (**Methods**).

We hypothesized that estimated liability is highly correlated with the true disease liability at the baseline age, thus can be used to study the age-dependent architecture of disease liability. We confirmed that the estimated liability achieves high accuracy across nine diseases, with an average liability-scale correlation of 0.603 (s.d. = 0.206 across nine diseases, **Supplementary Table 10**) with case-control status. We applied the same methods as in the quantitative trait analysis to the estimated liabilities to estimate *Δh*^2^ and *ρ*(*G*_*younger*_, *G*_*older*_). We validated that *h*^2^ of estimated liability is similar to the liability-scale *h*^2^of the respective diseases (**Supplementary Figure 7**). We found the heritability of the predicted liability significantly decreases for 5 of the 9 predicted disease liabilities (**Figure 3A**, **Supplementary Table 11**, mean *Δh*^2^ = −18.1%±2.7% for every 10 years of age), which supports the EA model of disease liability. We observe genetic correlations across age bins (*ρ*(*G*_*younger*_, *G*_*older*_)) are significantly different from 1 for coronary artery disease (CAD) and T2D (**Figure 3B**, **Supplementary Table 12**), which aligns with previous observations of heterogeneous age profiles in CAD^8^ and subtypes of T2D^23–25^. Taken together, we determined the estimated liabilities of asthma, hypertension, high lipid, COPD, and high cholesterol follow the EA model.

**Figure 3.**
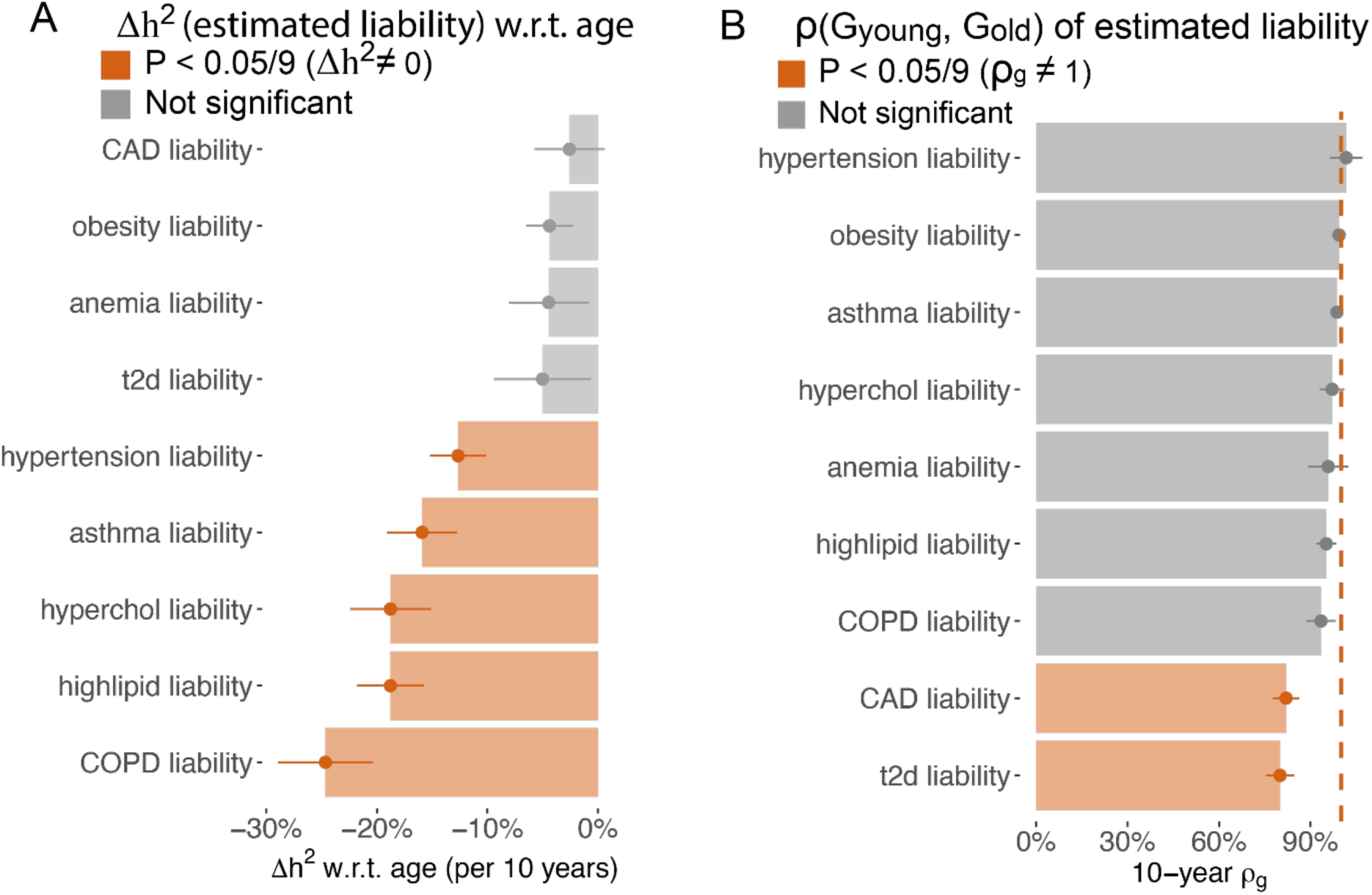
Age-dependent genetic architecture of estimated liability supports the EA model. (A) 10-year heritability change for estimated liability of 9 diseases; error bars show standard error via Monte-Carlo sampling. (B) 10-year genetic correlation for the estimated liability of 9 diseases. Error bars are standard error via meta-analysis of 10 age bin pairs. Numerical results are provided in **Supplementary Table 11-12**.

### Simulations implicate factors contributing to age-dependent disease PRS accuracy

We hypothesized that the following 3 factors could contribute to age-dependent disease PRS accuracy: (i) age-dependent liability under a liability threshold model^8,16^; (ii) prediction of incident cases (instead of prevalent cases)^26^; and (iii) exposure accumulation under our new EA model. We refer to these three hypotheses as factors (i) - (iii).

First, the intuition of factor (i) is that older cases have disease liability closer to healthy individuals compared to younger cases (see **Methods** and **Supplementary Note**). Under the liability-threshold model (**equation 1**), *L* increases as the population ages, which causes older cases to have liability that is closer to the healthy controls compared to the cases that reached the threshold at a younger age (**Figure 4A**, ref ^18^). Therefore, the difference between cases and controls becomes smaller for the older population, which causes decreasing prediction accuracy. In **Supplementary Note**, we show that the difference between the genetic values of cases and controls strictly decreases with age, which aligns with the intuition that liability of older cases is closer to healthy controls.

**Figure 4.**
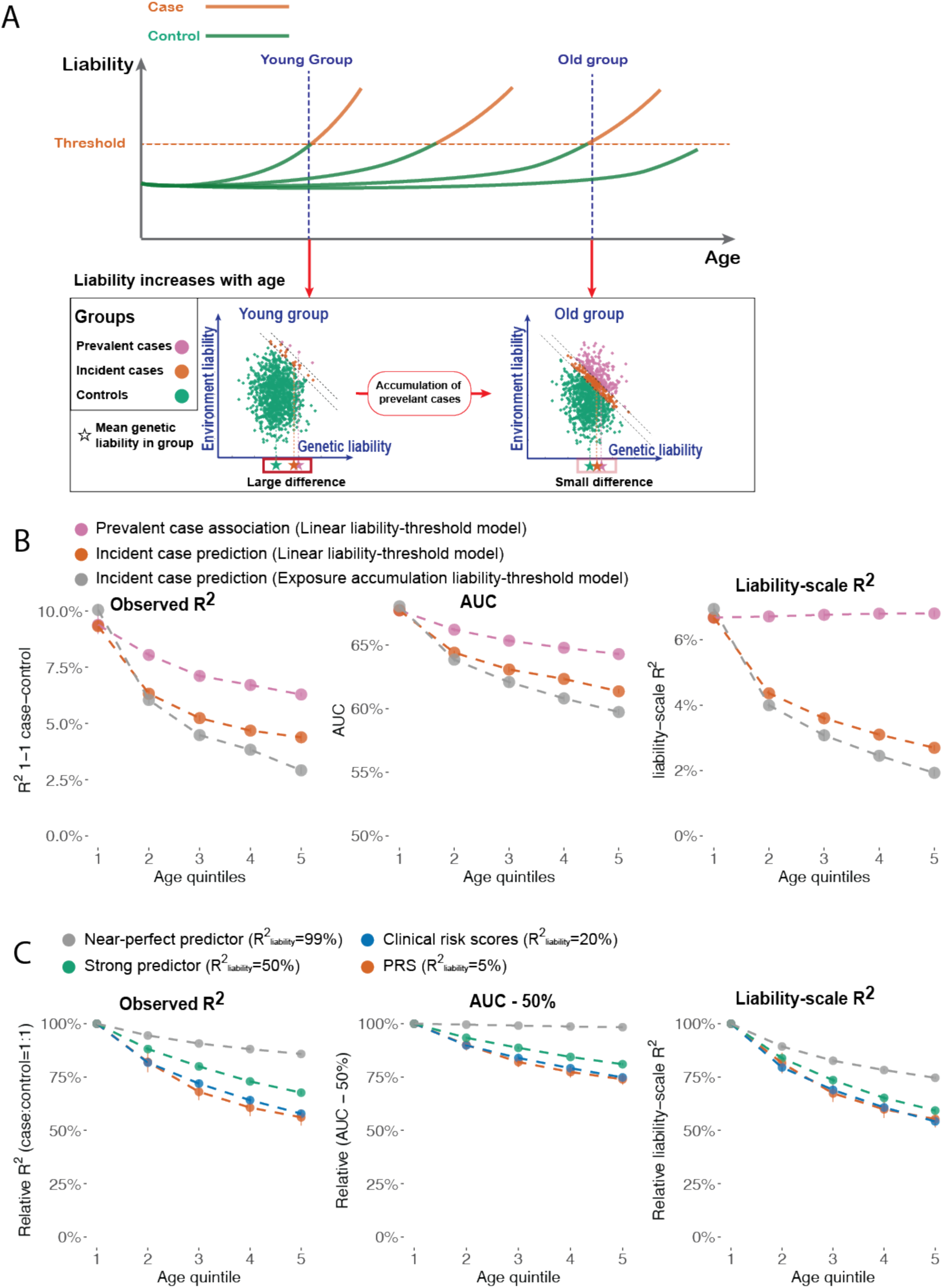
Liability-threshold model and the EA model explains age-dependent disease prediction accuracy in simulations. (A) Schematic figure explaining the decreasing disease prediction accuracy with age. In the top panel, each line represents the liability trajectory of an individual. The panel below shows the joint distribution of genetic and environmental liability at two age points. At later age points, cases are more similar to controls compared to earlier age points. (B) Prevalent case association accuracy (purple), incident case prediction accuracy (red), and Incident case prediction under the EA model (grey) across different age quintiles of cases. Prediction accuracy is measured by observed-scale R^2^ in 1:1 case-control matched samples, AUC, and liability scale R^2^ (C) Incident case prediction accuracy of predictors with different liability-scale R^2^.

Second, the intuition of factor (ii) is that excluding prevalent cases at baseline will create negative correlation between the predictable component and non-predictable component of disease liability, which reduces prediction accuracy (see **Supplementary Note)**. We note that there are two ways to define cases and controls at a specific age point (**Figure 4A**). First, *prevalent case association* defines cases as individuals who are diagnosed with the disease before the age point and controls as individuals who have not had a diagnosis of the disease at least until the age point; prevalent case association is a common choice in constructing genetic risk scores^20,27–30^. Second, *incident case prediction* defines cases as individuals who are diagnosed for the first time within a time window after the age point and controls as individuals who are not diagnosed at least until the end of the time window; incident case prediction is a common choice in clinical risk prediction^31,32^. We report results from both case-control definitions but focus on the latter because incident case prediction has more clinical utility.

Third, the intuition of factor (iii) is that PRS prediction accuracy decreases with age when heritability decreases with age (see **Supplementary Note**). Under the proposed EA model, heritability will decrease due to exposure accumulation, causing decreasing PRS accuracy with age.

We performed simulations to evaluate the contribution of factors (i)-(iii) to age-dependent prediction accuracy. Our simulation framework modeled liability increasing linearly with age. The age of onset is determined when the liability of an individual reaches a fixed threshold. We evaluate prediction accuracy using the observed *R*^2^ with 1:1 case-control ratio (*R*^2^_*obs*_), area under the ROC Curve (AUC), liability-scale *R*^2^ (*R*^2^_*liab*_), and log odds ratio (*logOR*). Further details are provided in the **Methods** section.

*R^2^*_*obs*_, AUC, and *logOR* all decrease with age for both prevalent case association and incident case prediction, while *R^2^*_*liab*_ decreases with age for incident case prediction but not for prevalent case association, supporting the contribution of factor (i) to age-dependent prediction accuracy (orange and purple curves in **Figure 4B; Supplementary Figure 8A**). Incident case prediction accuracy decreases more than the prevalent case association accuracy, supporting the contribution of factor (ii). We found that prediction accuracy under the EA model decreases more than under linear liability-threshold model, supporting the contribution of factor (iii) (grey curves in **Figure 4B)**. We estimated *R^2^*_*liab*_ under a normal distribution assumption^33^ in empirical data analyses. We evaluated the accuracy of this approximation using simulations and found it was accurate (**Supplementary Figure 8B**).

We show in the **Supplementary Note** that the *relative* contribution of factor (i) to age-dependent prediction accuracy depends on the *R^2^*_*liab*_ of the predictor to the underlying *L*. To validate this hypothesis, we simulated predictors with varying *R^2^*_*liab*_ to the underlying *L*, representing a perfect predictor (*R^2^*_*liab*_ = *99%*), a strong predictor (*R^2^*_*liab*_ = *50%*), a clinical score (*R^2^*_*liab*_ = *20%*), and a PRS (*R^2^*_*liab*_ = *5%*). Prediction accuracy decreased more steeply with age for weaker predictors, while the difference is negligible between the clinical risk score and PRS (**Figure 4C, Supplementary Figure 8A**).

Our analyses show that prediction accuracy decreases with age under the liability-threshold model (factor (i)) using both analytical results and simulations, while the decreasing slope is steeper when predicting incident cases (factor (ii)), predicting phenotypes that follow the EA model (factor (iii)), and the predictors have low overall *R^2^*_*liab*_.

### Age-dependent prediction accuracy of PRS is partially consistent with liability-threshold model and prediction of incident diseases

We next evaluated whether the age-dependent disease PRS accuracy is consistent with factor (i) and (ii) in empirical data. We computed PRS using BOLT-REML with fivefold cross validation for 9 diseases without age stratification. We divided the cases into age quintiles based on the earliest diagnosis age of each patient and created sample sets with a 1:1 case-control ratio to ensure each age bin has the same sample size and case-control ratio. We computed 4 metrics to measure incident case prediction accuracy: log hazard ratio (*logHR*), log odds ratio (*logOR*), the effect size of PRS in a linear regression on case-control status, and *R^2^*_*obs*_. We also compute

*R^2^*_*obs*_ for prevalent case association using the median age of each age quintile as the age index. We report the change of the 4 metrics (*logHR*, *logOR*, linear regression effects, and *R^2^*_*obs*_) by fitting a model where the metric changes linearly with age. Briefly, we test for non-zero changes in each metric with a likelihood ratio test between the model with age term and a null model where the metric is constant with age (**Methods**). We report the 10-year change of each metric relative to the estimates in the entire population.

We found PRS accuracy decreases with age across all diseases we studied, consistent with factor (i). We started by evaluating the *logHR* of each PRS to account for censoring. We found that the *logHR* of each PRS significantly decreased with age for 6 of the 9 diseases (p<0.05/9, **Figure 5A**, **Supplementary Table 10**), which aligns with a previous study that used lead GWAS associations and observed decreasing effect sizes with age^8^. On average, the *logHR* of each PRS in the oldest quintile was 51%±5% of the youngest quintile. For example, the *logHR* of the PRS for predicting hypertension was 0.39±0.01 for the youngest quintile of cases (median(age_onset_) = 47.1) and 0.16±0.01 for the oldest quintile of cases (median(age_onset_) = 72.2). The *logHR* of the PRS for predicting T2D is 0.32±0.02 for the youngest quintile of cases (median(age_onset_) = 47.7) and 0.22±0.02 for the oldest quintile of cases (median(age_onset_) = 72.8). These observations were consistent across different prediction accuracy metrics of PRS (*R^2^*_*obs*_, *logOR*, and linear regression effects, **Supplementary Figure 9, Supplementary Figure 10**, and **Supplementary Table 13**). We focus on *R^2^*_*obs*_ in later sections because it is on the same scale as **h**^2^. We found the average 10-year decrease for incident case prediction accuracy (*R^2^*_*obs*_) was greater than the 10-year decrease in accuracy for prevalent case association (**Figure 5B, Supplementary Table 14**), consistent with simulations of factor (ii) in **Figure 4B** (orange curves versus purple curves, respectively). For example, *R^2^*_*obs*_ for PRS predicting hypertension decreases 50.0%±1.7% every 10 years of age in incident case prediction and 26.8%±3.3% for prevalent case association. *R^2^*_*obs*_ for PRS predicting T2D decreases 25.1%±4.2% every 10 years of age in incident case prediction and no longer decreases for prevalent case association (slope= −2.9%±4.2% per 10 years). Our analyses indicate that PRS prediction accuracy generally decreases significantly with age.

**Figure 5:**
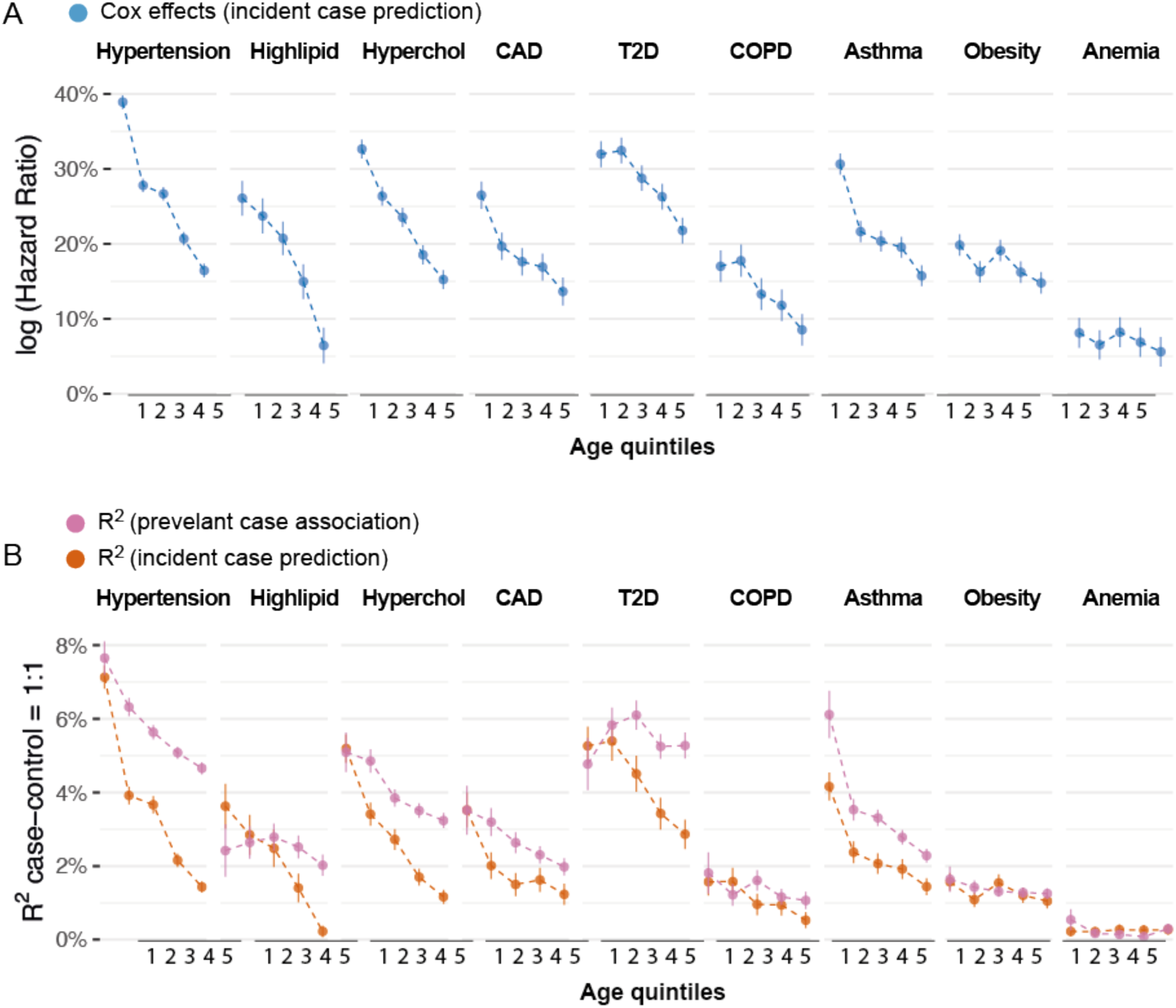
Age-dependent incident case prediction accuracy in 9 diseases. (A) Cox log hazard ratio of PRS on 9 diseases across 5 age quintiles. Each age bin has the same number of cases and 1:1 case-control ratio. Bars show standard error. (B) Age-dependent incident case prediction compared to prevalent case association accuracy in 9 diseases. Numerical results are provided in **Supplementary Table 13-14**.

### Exposure accumulation substantially impacts age-dependent disease PRS accuracy

We next evaluated whether the liability-threshold model (factor (i)), for incident case prediction (factor (ii)), and the EA model (factor (iii)) can explain age-dependent disease PRS accuracy in empirical data. We focus on incident case prediction so that all reported age-dependent prediction accuracy contains the contribution of factor (ii). We create quantitative risk scores (QRS) using quantitative traits to predict diseases. Because quantitative traits contain both genetic and non-genetic information, QRS does not contain the impact of the EA model. Therefore, age-dependent QRS prediction accuracy represents the contribution from factor (i) and factor (ii). We used the same 36 baseline quantitative traits to create QRS for each disease and add Gaussian noise so that QRS and PRS have the same *R^2^*_*liab*_ in the entire population (**Methods**). We compute the proportion of observed decrease in PRS accuracy with age that can be explained by each model to evaluate the contribution of the factor (i)-(iii) (**Methods**).

We found the liability-threshold model (factor (i)) and EA models (factor (iii)) together explain the observed age-dependent prediction accuracy of PRS for incident case prediction (factor (ii)). First, under the liability-threshold model for incident case prediction (factor (i)+(ii)), predictors using genetic data and predictors using non-genetic data (*i.e.* measurements of quantitative traits and environmental exposures) are expected to have the same age-dependent prediction accuracy, if they explain the same amount of variance in the liability ( *R^2^*_*liab*_) (**Figure 4C**). We used the change of QRS *R^2^*_*obs*_ with age to estimate the contribution from factor (i) + (ii) (**Methods**). We found 65.1%±11.3% of the change of PRS *R^2^*_*obs*_ with age are explained by these two factors (**Figure 6A**, **Supplementary Table 15** and **Supplementary Table 16**). The change of QRS *R^2^*_*obs*_ with age is significantly different from the change of PRS *R^2^*_*obs*_ with age for hypertension (*p*=6e-8), asthma (*p* = 9e-13), and CAD (*p* = 7e-7). We determined that factor (i) + (ii) explains most, but not all of the decreasing PRS prediction accuracy. Second, under the EA model (factor (iii)), the heritability of disease liability decreases with age, which causes PRS (which only captures genetic variance) accuracy to decrease with age. We found *Δh*^2^ of disease liability is not large enough to fully explain the decrease in PRS prediction accuracy (**Figure 6B**), suggesting the EA model in disease liability (factor (ii)) is insufficient to explain all of decreasing PRS prediction accuracy.

**Figure 6:**
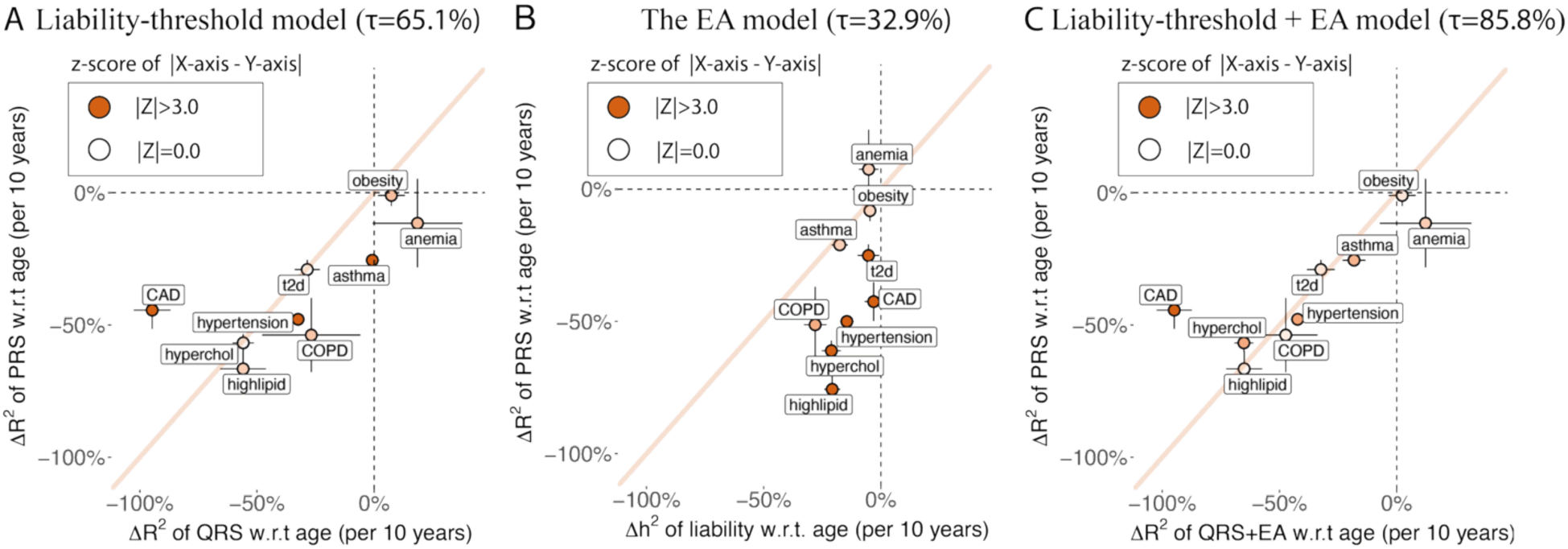
Liability-threshold model and the EA model explain age-dependent PRS prediction accuracy. (A) 10-year change in QRS prediction R^2^ under the liability-threshold model versus 10-year change in PRS prediction R^2^ for 9 disease traits. (B) 10-year heritability change in estimated disease liability versus 10-year change in PRS prediction R^2^ for corresponding disease traits. (C) 10-year change in QRS prediction R^2^ under the liability-threshold+EA model versus 10-year change in PRS prediction R^2^ for 9 disease traits. Diagonal line has slope equals to 1. Bars show the se and color shows the z-score of difference in X and Y values of each dot. *τ* is the proportion of observed decrease in PRS accuracy explained by each model (**Methods**). Numerical results are provided in **Supplementary Table 15-17**. We note that no estimates are significantly outside the plausible range and that allowing estimates outside of the plausible range is necessary to ensure unbiasedness of estimates.

Lastly, we combine the effect of EA model with liability-threshold model by computing the product of QRS *R^2^*_*obs*_ and **h**^2^ of predicted liability as the expected prediction accuracy of PRS, which captures the contribution of factors (i), (ii), and (iii). We determined that 85.8%±9.8% of the observed age profiles of PRS is explained by the factor (i)+(ii)+(iii) (**Figure 6C, Supplementary Table 17**). A model with factors (i)+(ii)+(iii) explains significantly more observed Δ*R*^2^_*obs*_(*PRS*) with age than the model without factor (iii) (*p* = 4e-6). We note that CAD cannot be explained by our models (*p* = 3e-7 for x and y difference in **Figure 6C**). CAD also has *ρ*(*G*_*younger*_, *G*_*older*_) < 1.0 (**Figure 3B**), suggesting that age-dependent genetic heterogeneity might cause the deviation from the liability-threshold + EA model. Taken together, our results suggest that the liability-threshold (factor (i)), incident case prediction (factor(ii)), and the EA model (factor (iii)) explain age-dependent prediction accuracy of PRS.

### Age-dependent accuracy of non-genetic predictors of disease

We investigated whether the liability-threshold model applies to age-dependent accuracy of both genetic and non-genetic predictors. We constructed risk predictors using the 36 quantitative measurements as non-genetic variables to further validate the liability-threshold model as a general model of disease. We note these 36 quantitative measurements contain both genetic and non-genetic (but predominantly non-genetic), but we use “non-genetic” to distinguish from analysis that focus on G. For each disease, we constructed 4 predictors with varying prediction power: (1) “QRS” predictor which is a LASSO predictor that uses all 36 quantitative measurements, (2) A “Top 3” predictor that is a joint linear predictor trained using the top three variables ranked by the *R^2^*_*liab*_ with target disease, (3) “single (strong)” predictors which are the quantitative measurements that rank 1st-3rd by the *R^2^*_*liab*_ with target disease, (4) “single (weak)” predictors which are the quantitative measurements that rank 4th and 5th. We trained all predictors using five-fold cross validation and estimated their prediction accuracy of diseases in each age quintile based on baseline UK Biobank visit. We then computed the change of *R^2^*_*obs*_ w.r.t. age for each predictor using the same method as the previous sections.

Predictors with greater prediction power in the entire population have less age-dependent prediction accuracy (**Figure 7A**, **Supplementary Table 18,** and **Supplementary Table 19**), as predicted by simulation results (**Figure 4C, Supplementary Note**). Relative change of 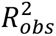 w.r.t. age is −47.6%±1.3% for “single (weak)” predictors (avg(*R^2^*_*liab*_) = 9.8%) and −23.6%±1.1% for “QRS” predictors (avg(*R^2^*_*liab*_) = 32.3%), averaging across the 9 diseases. We further expand to 10 predictors for each disease (QRS, “Top 3”, each of the top 5 predictors, “top 2”, PRS, PRS + QRS, restricting to predictors with *R^2^*_*liab*_ > 5% for adequate power) and found strong positive correlations between relative change of 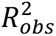 w.r.t. age and *R^2^*_*liab*_ across disease-predictor pairs (**Figure 7B**, slope = 0.64, *p* =1.2e-5). We conclude that predictors that use environment variables have decreasing accuracy with age and the effect is stronger for predictors with low overall prediction accuracy, suggesting combining genetic and non-genetic predictors can increase prediction power in older populations.

**Figure 7.**
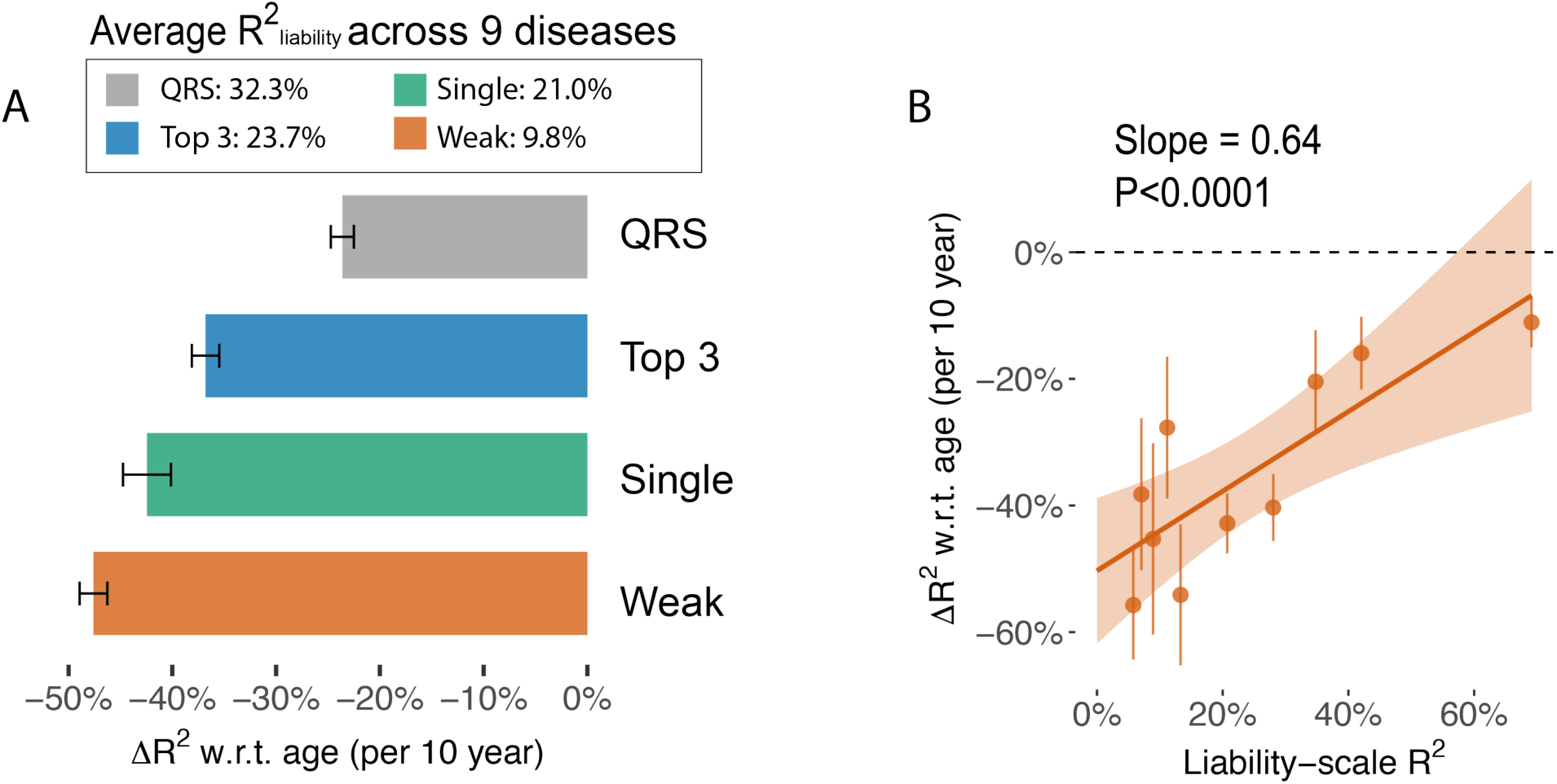
Risk models with higher liability-scale R^2^ are less attenuated when predicting late-onset diseases. (A) Age-dependent prediction accuracy of predictors averaged across 9 diseases. X-axis shows the per-10-year change of prediction R^2^ computed in 1-1 case control matching groups. QRS: predicted liability using 36 quantitative traits. Top 3: linear predictors use the top three quantitative traits that have the highest correlation with the diseases. Single (Strong): using a single quantitative trait to predict incident diseases, averaged across the top three traits as in the “Top 3” group; Single (Weak): the average between 4th and 5th predictor of each disease. (B) Scatter plot comparing the liability-scale R^2^ (without age stratification) with the change of R^2^ w.r.t. age across predictor-disease pairs. Predictor-disease pairs are grouped into 10 deciles based on liability-scale R^2^ for visualization. The red line shows a regression line with confidence interval. Numerical results are provided in **Supplementary Table 18-19**.

## Discussion

We systematically quantified age-profiles of genetic and environmental variation to understand how complex disease architecture and polygenic risk score (PRS) accuracy vary with age. We found exposure accumulation is common in quantitative traits and complex diseases. We found the age-dependent PRS accuracy can be explained by a combination of a liability-threshold model and the exposure accumulation model. Our analysis was enabled by a novel approach to study estimated disease liability. Finally, we found both genetic and non-genetic predictors decrease in prediction accuracy with age in empirical data and simulations.

Our study represents an advance over previous studies that investigate age-dependent effects. First, we identify changes in genetic and environmental variance across 16 complex quantitative traits and 9 diseases. While previous work has studied age-dependent effects, they have focused on single variant effects^5^ or have studied a limited set of phenotypes^4^. Second, to the best of our knowledge, our work is the first to quantify how environmental variance in complex traits accumulates with age using longitudinal data. Third, our work is the first to provide a quantitative explanation for the decrease in prediction accuracy for genetic and non-genetic predictors. While other studies have observed this phenomenon, they have not been able to explain it^8,13^. We note, however, that CAD was an exception to the EA model and, while we speculate that this may be due to age-dependent genetic heterogeneity, this is something of a mystery to be resolved in subsequent studies. Lastly, our work demonstrates that PRS trained and tested using prevalent case association^20,27–30^ shows different age-dependent accuracy compared with incident case prediction, underscoring the importance of evaluating PRS accuracy using incident case prediction, which has greater clinical utility^34–37^.

Our study has several implications. First, the EA model suggest GWAS study can increase power by accounting for age-dependent heritability. In particular, we hypothesize that mixed effect model can increase power by modelling age-specific random effects. Second, the EA model suggests when incorporating PRS in clinical risk models, PRS should be weighted differently with age to increase power. Third, our results suggest all predictors (*i.e.* genetic or non-genetic) of disease have age-dependent accuracy, while predictors with stronger prediction power in the entire population will have more robust prediction accuracy at older age. This motivates constructing predictors that combine multiple data modalities to achieve maximal prediction accuracy across age. Fourth, the observed exposure accumulation suggests genetic effects contribute more to phenotypic variation at early ages, suggesting ascertaining environment variables is critical to accurately predict phenotypes at later ages. Lastly, our results suggest recruiting younger cases can improve power for discovering both genetic and environment biomarkers for diseases. All of these implications motivate directions for future research.

Our study has several limitations. First, our modeling is not able to detect uniformly decreasing genetic variance which can cause decreasing *h*^2^ with age and *ρ*(*G*_*young*_, *G*_*old*_) close to 1. However, we believe this is not likely because we found evidence of accumulating environmental variance in the 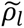 analyses which explains the decreasing *h*^2^ (**Figure 2C**). Second, the age range of our study sample is restricted, with baseline visit age between 40 to 79 years of age. Future analyses with a wider age distribution may discover stronger effects. Third, we use a cross-sectional analysis, which is susceptible to recruitment bias. We tested whether age-dependent recruitment bias of healthier individuals could explain decreasing heritability estimates with age and observed minimal heritability differences between the two groups, suggesting a minimal effect of biased recruitment. Fourth, our analyses uses only individuals of European individuals. While it is of interest to perform the analysis on individuals of other ancestries, currently available data of other genetic ancestry might not have sufficient power for age-stratified analyses. Fifth, our analyses use the UK Biobank data set. It is of interest to replicate the analysis in other cohorts such as All of Us^38^. Sixth, majority of our findings rely on the validity of liability-threshold model, although liability-threshold model has been widely accepted and used^16,17,39–45^. Despite these limitations, our work identifies exposure accumulation as a key aspect age-dependent disease architecture, and our results explain the decreasing genetic and non-genetic disease prediction accuracy with age.

## Methods

### Study population in UK Biobank

We used the disease diagnosis information from 180,026 individuals whose primary care data (General Practice, GP) was available and of British Isle ancestry. The primary care data are coded in readv2 which were mapped to SNOMED CT (https://digital.nhs.uk/services/terminology-and-classifications/read-codes), SNOMED CT codes were then mapped to ICD-10 codes (using NHS digital’s code mapping https://digital.nhs.uk/), which were combined with ICD-10 codes from Hospital Episode Statistics (HES) and mapped to PheCodes ^46^. We extracted the earliest diagnosis age using the earliest records from the combined primary care and HES data, which are used as age labels in our analysis. The choice of focusing on individuals with both primary data and HES data was to get the best age at diagnosis for each disease. When combining GP and HES data to extract diagnosis, correlation between binary and quantitative traits increased compared to only using HES data to extract disease (**Supplementary Figure 11**), suggesting that combining GP and HES data capture diagnoses more accurately as correlation decreases when variables are measured with noise.

### Selection of quantitative and binary traits

We extracted 36 quantitative traits measured at baseline visit of UK Biobank based on ref. ^20^. As our goal is to study disease liability, we selected 9 diseases using phecodes that have interpretable underlying quantitative traits from the 36 quantitative traits (e.g. Hb1Ac for T2D, BMI for obesity, LDLc for hypercholesterolemia) and prevalence higher than 5% (**Supplementary Table 3**). Disease status and age-at-diagnosis were extracted from GP+HES data described in the previous section.

We corrected blood biochemistry measurement for cholesterol, hypertensive, and diabetic medications: cholesterol (+1 s.d.), LDL-c (+ 1 s.d.), and triglyceride levels (+0.5 s.d.) were corrected for lipid lowering medication^47^; systolic blood pressure (+15 mm Hg) and diastolic blood pressure (+ 10 mm Hg) were corrected for hypertension medication^48^; HbA1c level is corrected for non-insulin diabetic medication (+ 10.929) and insulin drugs (+ 1.3 x 10.929)^49^. The respective changes for individuals who are taking the medication are in the parentheses after each trait; s.d. is the standard deviation computed in the entire population. We verified that the correlation between the corrected quantitative traits and corresponding binary traits is higher compared to the correlation between uncorrected quantitative traits and the same set of binary traits. *ρ*(*LDL*, *hypercholesterolemia*): 0.188 post-medication correction vs −0.005 pre-correction. *ρ*(*Systolic blood pressure*, *hypertension*): 0.519 post-medication correction vs 0.425 pre-correction. *ρ*(*HbA*1*c*, *T*2*D*): 0.565 post-medication correction vs 0.524 pre-correction. Both corrected and uncorrected values were included in the final 36 quantitative traits.

In our analyses, smoking status is defined as an ordinal variable with the following values: never (0), previous (1), current (2) (**Supplementary Table 4**).

To select quantitative traits that are most relevant to the nine diseases, we further computed the correlation between the 36 quantitative traits and the nine disease traits and selected 16 of the 36 quantitative traits based on liability-scale correlation ^33^; the included quantitative traits are among the top 3 correlated traits for at least one of the nine binary diseases.

### Heritability and genetic correlation

We estimated heritability using the REML function of BOLT-LMM^19^ version 2.4.1. 180,026 individuals were divided into 5 age bins based on the baseline visit age. We performed REML analysis for each age bin using sex and baseline visit age as covariates; we use the same 536,492 HapMap3 SNPs that have moderate LD with each other which is the same as the PRS section, see below. We also computed the heritability of 16 quantitative traits and nine diseases in the entire population.

We estimated genetic correlation between age bins for the 16 quantitative traits using LDscore regression^50^. First, we used the mixed-effect model to compute summary statistics for each age bin using BOLT-LMM version 2.4.1. We used the same set of variants and individuals as the heritability estimation. Since BOLT-LMM is a mixed effect model that controls population stratification, we constrained the heritability intercept of LDSC to 1.0; we constrained covariance intercept to be 0 as there is no sample overlap between different age bins. We run cross-trait LDSC for each age-bin pair (10 pairs in total for 5 age bins) to estimate the genetic correlation and its standard error.

### Testing for heritability changes across age bins

We tested the age-dependency of heritability using heritability estimates with standard error across the five age bins. We used likelihood ratio tests to identify non-constant heritability across age bins. We tested two nested models: *H*_0_: *h*^2^∼*N*(0 × *age* + *b*, *s*. *e*. (*h*^2^)) and *H*_1_: *h*^2^∼*N*(*a* × *age* + *b*, *s*. *e*. (*h*^2^)). We estimated *b* under *H*_0_ and *a*, *b* under *H*_1_ using maximum likelihood estimators across the five heritability estimates. For each age quintile, we use the median age to fit the model. The slope *a* estimated under *H*_1_ are the per year change of *h*^2^. We use Monte-Carlo sampling of *h*^2^ for each age quintile from a Gaussian distribution with mean equals to the point estimates and variance equals to s.e.. We estimate *a*, *b* of each Monte-Carlo sample. The s.d. of *a* estimates across Monte-Carlo samples are the s.e. of the estimates. We note this approach relies on the fact that samples are non overlapping across age quintiles.

In all analyses, we report the relative change of heritability compared to the heritability estimate in the entire population.

### Testing for genetic correlation less than 1 between age bins

The quantity of interest is the difference between genetic correlation and 1: 1 − *ρ_g_*. First, we scale genetic correlation using *ρ*_*normalised*_ = *exp*(10 ×*ln ρ*_*Δage*_/ *Δage*) to account for the varying age differences of age-bin pairs; the age differences are computed using the median of each age quintile. Scaling on the exponential scale aligns with a Wiener process of liability w.r.t age (see below). For 5 age bins, we can estimate 10 genetic correlations between pairs of age bins; these 10 genetic correlations have four degrees of freedom. Therefore, we use inverse-variance weighting to meta-analyse four of the 10 scaled *ρ_g_* (we use *ρ_g_* of neighboring age-bin pairs 1-2, 2-3, 3-4, and 4-5) to compute one 10-year *r_g_* for each trait; standard errors of the 10-year *r_g_* estimates are used for inverse-variance weights.

### Computing phenotypic correlation across age points

Under the assumption that random variance accumulates linearly w.r.t. age (as in a Wiener process), the correlation between liability across age bins is a function on the multiplicity scale *ρ*_*Δage*_ = *e*^−*β*^ ^*Δage*^. Therefore, we computed the expected 10-year genetic and phenotypic correlation by *exp*(10 ×*ln ρ*_*Δage*_/ *Δage*) to make the correlations comparable for age bin pairs with different age differences. We computed phenotype correlations across individuals who have three visits to the UK Biobank. We remove the impact of measurement error by computing a ratio of two correlations *ρ*_*Δage*_ = *ρ*_*l*_(*t*_1_, *t*_3_)/*ρ*_*l*_ (*t*_1_, *t*_2_), which cancels out the measurement error, and we use the average age difference between second and third visit (*Δage* = *t*_3_ − *t*_2_) to scale the estimate.

### Testing the impact of selection in recruitment process

To test for whether the age-dependent heritability changes can be attributed to selected recruitment of healthier individuals at older age groups^21^, we computed the number of past diagnosis records as an indicator of health status and created samples with matched age but different past diagnosis. We used health records from both GP and HES, and divided the target population into two groups, “healthy” versus “unhealthy”, using the median number of prevalent records (median=16). We then use inverse probability weights to sample 45,267 individuals from both groups, resulting in two samples that have matched age distributions (mean: 55.9 vs. 56.6, respectively) but different numbers of past diseases (mean: 6.8 vs 34.6, respectively); in comparison, top and bottom age quintile has an average age of 67.5 and 45.3 and an average of 24.2 and 18.7 of past diseases, respectively. We compute heritability of the 16 quantitative for both groups using the same procedure as for age quintiles.

### PRS construction

We obtained 536,492 HapMap3 SNPs using pruning parameters of window size=50 kb, step size=5 kb, and r^2^ threshold=0.8; selection of SNPs makes mixed effect model computational tractable. We then used BOLT-LMM to estimate PRS, using genetic sex and age at baseline as covariates, using 5-fold cross validation for the 9 diseases and 16 quantitative traits. PRS are scaled to have mean=0 and sd=1 in the entire population.

### PRS incident case prediction and prevalent case association

For incident case prediction, we first divided cases into five age quintiles based on their first age-at-diagnosis; for each age quintile, we define case control status based using interval-censor procedure ^8^, where cases are defined as those who are diagnosed within the age interval and controls are defined as those who remained healthy till the end of age interval or lost to follow up within the age interval. We then down sampled the controls for each age bin to have the same number as the cases. As a result, for a given disease, each age interval has exactly the same number of cases and controls which ensures the odds ratio and hazard ratio are comparable across each age bin. When evaluating PRS using linear, logistic, and Cox regression models, we included genetic sex and top five genetic PCs as covariates. When evaluating PRS using observed-scale R^2^, we first regress the outcome on genetic sex and top five genetic PCs, then compute Pearson’s correlation between the residual and PRS; the square of Pearson’s correlation is the observed-scale R^2^ with 1:1 case control ratio.

For prevalent cases association, we used the five median case age of the respective age intervals as the baseline age points and considered all individuals who have been followed up past the baseline age point. The prevalence cases are defined as those who have at least one diagnosis record before the baseline age point. We sampled controls to be the same number as cases for each age point, so that for a given disease, the effect sizes of linear, logistic, and Cox regression across five baseline age points are comparable. The regression models and observed-scale R^2^ were computed using the same covariates as in incident case association.

### Estimated disease liability

We use 36 continuous traits measured at baseline to predict the 9 binary disease traits; the resulting predictor is the *estimated disease liability*. Cases are defined as the union of both incident and prevalent disease. We use LASSO linear model (instead of logistic regression as we assume a liability-threshold model) to train the prediction models with 10-fold cross validation; the regularization coefficients are chosen by the cross-validation with both 1se (default setting of the R glmnet package^51,52^) and 2se (stronger regularization). We chose the predicted liability using the 2se model as it has a similar liability-scale correlation (average=0.60 across 9 diseases) with the binary trait to the 1se model (average=0.61 across 9 diseases), while being more explainable and less susceptible to overfitting. The models were fitted in the entire population without including age as a covariate so that the predicted disease liability represents the approximate disease liability at the age of baseline visit.

We used the same procedure as the quantitative traits analysis to estimate heritability and genetic correlation for predicted liability. We used BOLT-REML to estimate heritability. We used BOLT-LMM to compute GWAS summary statistics then use cross-trait LDSC to estimate genetic correlation of each age-bin pair (10 pairs in total for 5 age bins) to estimate the genetic correlation and its standard error. We did not constrain intercepts for cross-trait LDSC to avoid potential bias population relatedness.

### Simulation of linear liability-threshold model

We simulated genetic variance G and environment variance E from normal distribution with mean 0 and variance equal to *h*^2^ and (1−*h*^2^) respectively. Liability *L* equals the sum of *G* and *E* for each individual. To simulate liability increasing with age, we take 2% of the simulated individuals with the highest liability as the youngest disease group, top 2-4% of liability as the second age group and repeated the procedure to obtain five groups of individuals with increasing onset age. Each disease threshold corresponds to the disease cut off at an age point. Prevalent cases are defined as the individuals that have liabilities higher than the threshold at a specific age point; incident cases are defined as individuals in the bin near the liability threshold at a specific age point (i.e. all individuals with liability near the corresponding liability); controls are defined as all individuals excluding prevalent cases (i.e. all individuals have liability lower than the corresponding liability).We simulated 100,000 individuals, with *h*^2^=0.3 and using five age bins of 2% of individuals, which matches the disease prevalence and sample size in empirical analyses.

We note our simulation uses percentiles of individuals to represent cases with increasing age, which aligns with the empirical analysis (see below) where we divided cases into quintiles based on the diagnosis age. We note under the proportional amplifciation model, the liability percentile of each individual does not change, which means this model is invariant to proportional amplification.

### Simulation of EA liability-threshold model

We simulated initial liability L using the same procedure as the linear liability-threshold model. For each age bin, we sample cumulative environmental variance *E*_*a*_ from normal distribution with 0 mean and 0.05 variance. We then cumulatively add these sampled environmental variances to the liability when moving to an older age quintile, which will result in 20% more accumulated variance in the fifth age quintile. In empirical data, we found ∼20% decrease in heritability over 20 years for quantitative traits, which motivates our choice of simulation parameters.

We defined prevalent cases as individuals who have got disease at any prior age bin or the current age bin, and incident cases as those who get disease within the current age bin. For each age bin, we take the top 2% individuals who have not prevalent diseases to simulate that liability increases with age. We note this simulation procedure makes the number of cases equal to previous simulation of the linear model so the metrics in **Figure 4B** are comparable.

### Estimating age-dependent prediction accuracy under the linear liability-threshold model

We use rank normal transformation of estimated liability to create a *quantitative risk score (QRS)*. QRS combines both G and E information which is different from PRS that only captures G information; under the linear liability-threshold model, QRS and PRS has the sample age-dependent prediction accuracy profile when their liability-scale R^2^ are equivalent across the population (**Figure 4C**). We performed the following 6 steps to use QRS to estimate expected age-dependency of PRS under the liability-threshold model. (1) QRS are rank normal transformed to have similar distributions as PRS. (2) QRS are adjusted for age at measurement and genetic sex; PRS are adjusted for first 5 genetic PCs and genetic sex. Adjustment of covariates ensures QRS and PRS are comparable under the linear liability-threshold model for each age bin, as QRS will change with age while PRS does not change with age. (3) We computed the liability-scale R^2^ of QRS and PRS using all individuals with primary care information (N = 215,162); cases are defined as the union of prevalent cases and incident cases. We computed observed-scale R^2^ for both QRS and PRS and transformed observed-scale R^2^ to liability-scale R^2^ using equation 14 of ref ^33^. (4) We added gaussian noise (mean = 0) to QRS so that the new QRS has the same liability-scale R^2^ as PRS. (5) We computed QRS observed-scale R^2^ for each age bin using the same case-control definition as incident case prediction of PRS. We note a caveat that some QRS are measured after disease diagnosis. Our choice is based on two reasons: first, under the linear liability-threshold model, QRS (age and sex adjusted) represents the liability that remains constant across age; second, observed-scale R^2^ for QRS and PRS are computed using the same samples, making them comparable. We use QRS to predict incident cases at baseline visit and found similar results, see sections pertains to “non-genetic predictors”. (6) We performed Monte-Carlo sampling to account for two sources of uncertainty: (i) the process to add noise to QRS and (ii) control sampling within each age bin. We reported mean and standard deviation of 50 Monte-Carlo sampling estimates for each parameter.

We define the proportion of observed decrease in PRS accuracy explained by each model as:

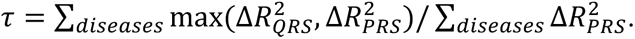

The max in the numerator reflects the constraint that the proportion of age-dependent PRS accuracy that can be explained is less than 100%. Therefore, diseases with more decrease in QRS accuracy than PRS accuracy are assigned the same value in numerator and denominator. This choice is justified by empirical data as we do not observe significantly more decrease in QRS accuracy than PRS accuracy at a nominal *p*-value of 0.05, except for CAD.

### Estimating age-dependent prediction accuracy under the EA + liability-threshold model

We multiplied the estimated slope from QRS with the slope of *h*^2^ of predicted liability to estimate the expected decrease under the EA + liability-threshold model. Under the EA model PRS prediction accuracy is proportional to *h*^2^ of disease liability at each age point. The EA + liability-threshold model estimates capture both the age-dependency from the linear liability-threshold model (captured by QRS) and the EA model (captured by the *Δh*^2^ of predicted liability). The expected change is computed as 1 - (1 + *ΔR*^2^_*QRS*_) ×(1 + *Δh*^2^_*predicted*–*liability*_). We use Monte-Carlo sampling of *ΔR*^2^_*QRS*_ and *Δh*^2^_*predicted*–*liability*_ to estimate the standard error of the estimator; we omit the sampling covariance between *ΔR*^2^_*QRS*_ and *Δh*^2^_*predicted*–*liability*_ as they were estimated from approximately independent information (*i.e. ΔR*^2^_*QRS*_ were estimated using age quintiles of cases and baseline measurements, while *Δh*^2^_*predicted*–*liability*_ uses genetic information).

### Incident case prediction with non-genetic predictors

For environment variable predictors, we divided incident cases into five age bins using the age at first diagnosis visit. We note that in contrast to PRS prediction accuracy, we excluded individuals who have prevalent cases before baseline visits as environment variables are measured at baseline visit. The definition of incident cases and controls for each age quintile, down sampling of control is the same as the PRS incident case prediction. We computed observed-scale R^2^ for each environment predictor by regression of sex and top five genetic PCs from the outcomes and computed Pearson’s correlation between the outcome residual and the predictor; the square of Pearson’s correlation is the observed-scale R^2^ with 1:1 case control ratio.

We analyzed 10 predictors including 9 environment predictors and PRS. We computed the relative change of observed-scale R^2^ compared to the observed-scale R^2^ estimate in the entire population. Observed-scale R^2^ in the entire population were transformed to liability-scale R^2^ using equation 14 of ref ^33^. When computing the relationship between liability-scale R^2^ (in the entire population) to the change of observed-scale R^2^, we only include 61 of 90 predictors that have liability-scale R^2^ larger than 5% to ensure there is sufficient power to detect age-dependency.

## Code Availability

Code to replicate all analyses in the manuscript is available at https://github.com/Xilin-Jiang/GxAge

## Data Availability

Summary data in the manuscript is available at https://github.com/Xilin-Jiang/GxAge

## Acknowledgements

Use of the UK Biobank data was performed under application number 7439. This research was funded by Wellcome Trust early-career award 227566/Z/23/Z (X.J.) and the National Institute Of General Medical Sciences of the National Institutes of Health under Award Number R35GM16046 (A.D.). M.I. is supported by the Munz Chair of Cardiovascular Prediction and Prevention and the NIHR Cambridge Biomedical Research Centre (NIHR203312) as well as by the UK Economic and Social Research 878 Council (ES/T013192/1). J.D. is supported by a British Heart Foundation Personal Chair. This work was supported by core funding from the British Heart Foundation (RG/F/23/110103), NIHR Cambridge Biomedical Research Centre (NIHR203312), BHF Chair Award (CH/12/2/29428), Cambridge BHF Centre of Research Excellence (RE/24/130011) and by Health Data Research UK, which is funded by the UK Medical Research Council, Engineering and Physical Sciences Research Council, Economic and Social Research Council, Department of Health and Social Care (England), Chief Scientist Office of the Scottish Government Health and Social Care Directorates, Health and Social Care Research and Development Division (Welsh Government), Public Health Agency (Northern Ireland), British Heart Foundation and the Wellcome Trust. The views expressed are those of the authors and not necessarily those of the NIHR, the Department of Health and Social Care, or the National Institutes of Health.

## References

1. Kroemer, G. et al. From geroscience to precision geromedicine: Understanding and managing aging. Cell 188, 2043–2062 (2025).

2. Moqri, M. et al. Biomarkers of aging for the identification and evaluation of longevity interventions. Cell 186, 3758–3775 (2023).

3. Simino, J. et al. Gene-age interactions in blood pressure regulation: a large-scale investigation with the CHARGE, Global BPgen, and ICBP Consortia. Am. J. Hum. Genet. 95, 24–38 (2014).

4. Robinson, M. R. et al. Genotype-covariate interaction effects and the heritability of adult body mass index. Nat. Genet. 49, 1174–1181 (2017).

5. Winkler, T. W. et al. Genetic-by-age interaction analyses on complex traits in UK Biobank and their potential to identify effects on longitudinal trait change. Genome Biol. 25, 300 (2024).

6. Miao, J. et al. PIGEON: a statistical framework for estimating gene-environment interaction for polygenic traits. Nat. Hum. Behav. 1–15 (2025).

7. Zhu, C. et al. Amplification is the primary mode of gene-by-sex interaction in complex human traits. Cell Genomics 3, (2023).

8. Jiang, X., Holmes, C. & McVean, G. The impact of age on genetic risk for common diseases. PLoS Genet. 17, e1009723 (2021).

9. Sakaue, S. et al. Trans-biobank analysis with 676,000 individuals elucidates the association of polygenic risk scores of complex traits with human lifespan. Nat. Med. 26, 542–548 (2020).

10. Jukarainen, S. et al. Genetic risk factors have a substantial impact on healthy life years. Nat. Med. 28, 1893–1901 (2022).

11. Dahl, A. et al. A Robust Method Uncovers Significant Context-Specific Heritability in Diverse Complex Traits. Am. J. Hum. Genet. 106, 71–91 (2020).

12. Pazokitoroudi, A. et al. A scalable and robust variance components method reveals insights into the architecture of gene-environment interactions underlying complex traits. Am. J. Hum. Genet. 111, 1462–1480 (2024).

13. Thompson, D. J. et al. A systematic evaluation of the performance and properties of the UK Biobank Polygenic Risk Score (PRS) Release. PLoS One 19, e0307270 (2024).

14. Argentieri, M. A. et al. Integrating the environmental and genetic architectures of aging and mortality. Nat. Med. 31, 1016–1025 (2025).

15. Durvasula, A. & Price, A. L. Distinct explanations underlie gene-environment interactions in the UK Biobank. Am. J. Hum. Genet. 112, 644–658 (2025).

16. Zaitlen, N. et al. Informed conditioning on clinical covariates increases power in case-control association studies. PLoS Genet. 8, e1003032 (2012).

17. Pedersen, E. M. et al. Accounting for age of onset and family history improves power in genome-wide association studies. Am. J. Hum. Genet. 109, 417–432 (2022).

18. Aschard, H., Zaitlen, N., Lindström, S. & Kraft, P. Variation in predictive ability of common genetic variants by established strata: the example of breast cancer and age. Epidemiology 26, 51–58 (2015).

19. Loh, P.-R. et al. Efficient Bayesian mixed-model analysis increases association power in large cohorts. Nat. Genet. 47, 284–290 (2015).

20. Weissbrod, O. et al. Leveraging fine-mapping and multipopulation training data to improve cross-population polygenic risk scores. Nat. Genet. 54, 450–458 (2022).

21. Fry, A. et al. Comparison of sociodemographic and health-related characteristics of UK Biobank participants with those of the general population. Am. J. Epidemiol. 186, 1026–1034 (2017).

22. Tibshirani, R. Regression shrinkage and selection via the lasso. Journal of the royal statistical society series b-methodological 58, 267–288 (1996).

23. Udler, M. S. et al. Type 2 diabetes genetic loci informed by multi-trait associations point to disease mechanisms and subtypes: A soft clustering analysis. PLoS Med. 15, e1002654 (2018).

24. Jiang, X. et al. Age-dependent topic modeling of comorbidities in UK Biobank identifies disease subtypes with differential genetic risk. Nat. Genet. 55, 1854–1865 (2023).

25. Suzuki, K. et al. Genetic drivers of heterogeneity in type 2 diabetes pathophysiology. Nature 1–11 (2024).

26. Celentano, D. D., Szklo, M. & Farag, Y. Gordis Epidemiology. (Elsevier - Health Sciences Division, Philadelphia, PA, 2024).

27. Khera, A. V. et al. Genome-wide polygenic scores for common diseases identify individuals with risk equivalent to monogenic mutations. Nat. Genet. 50, 1219–1224 (2018).

28. Ge, T., Chen, C.-Y., Ni, Y., Feng, Y.-C. A. & Smoller, J. W. Polygenic prediction via Bayesian regression and continuous shrinkage priors. Nat. Commun. 10, 1776 (2019).

29. Privé, F., Arbel, J. & Vilhjálmsson, B. J. LDpred2: better, faster, stronger. Bioinformatics 36, 5424–5431 (2021).

30. Zheng, Z. et al. Leveraging functional genomic annotations and genome coverage to improve polygenic prediction of complex traits within and between ancestries. Nat. Genet. 56, 767–777 (2024).

31. SCORE2 working group and ESC Cardiovascular risk collaboration. SCORE2 risk prediction algorithms: new models to estimate 10-year risk of cardiovascular disease in Europe. Eur. Heart J. 42, 2439–2454 (2021).

32. Patel, A. P. et al. A multi-ancestry polygenic risk score improves risk prediction for coronary artery disease. Nat. Med. 29, 1793–1803 (2023).

33. Lee, S. H., Goddard, M. E., Wray, N. R. & Visscher, P. M. A better coefficient of determination for genetic profile analysis. Genet. Epidemiol. 36, 214–224 (2012).

34. Chatterjee, N., Shi, J. & García-Closas, M. Developing and evaluating polygenic risk prediction models for stratified disease prevention. Nat. Rev. Genet. 17, 392–406 (2016).

35. Torkamani, A., Wineinger, N. E. & Topol, E. J. The personal and clinical utility of polygenic risk scores. Nat. Rev. Genet. 19, 581–590 (2018).

36. Kullo, I. J. et al. Polygenic scores in biomedical research. Nat. Rev. Genet. 23, 524–532 (2022).

37. Kullo, I. J. Promoting equity in polygenic risk assessment through global collaboration. Nat. Genet. 56, 1780–1787 (2024).

38. Genomic data in the All of Us Research Program. Nature 1–7 (2024).

39. Dempster, E. R. & Lerner, I. M. Heritability of threshold characters. Genetics 35, 212–236 (1950).

40. Wray, N. R. & Goddard, M. E. Multi-locus models of genetic risk of disease. Genome Med. 2, 10 (2010).

41. Lee, S. H., Wray, N. R., Goddard, M. E. & Visscher, P. M. Estimating missing heritability for disease from genome-wide association studies. Am. J. Hum. Genet. 88, 294–305 (2011).

42. Witte, J. S., Visscher, P. M. & Wray, N. R. The contribution of genetic variants to disease depends on the ruler. Nat. Rev. Genet. 15, 765–776 (2014).

43. Hujoel, M. L. A., Gazal, S., Loh, P.-R., Patterson, N. & Price, A. L. Liability threshold modeling of case-control status and family history of disease increases association power. Nat. Genet. 52, 541–547 (2020).

44. Hujoel, M. L. A., Loh, P.-R., Neale, B. M. & Price, A. L. Incorporating family history of disease improves polygenic risk scores in diverse populations. Cell Genom. 2, 100152 (2022).

45. Pedersen, E. M. et al. ADuLT: An efficient and robust time-to-event GWAS. Nat. Commun. 14, 5553 (2023).

46. Wu, P. et al. Mapping ICD-10 and ICD-10-CM Codes to Phecodes: Workflow Development and Initial Evaluation. JMIR Med Inform 7, e14325 (2019).

47. Kofink, D. et al. Statin Effects on Metabolic Profiles: Data From the PREVEND IT (Prevention of Renal and Vascular End-stage Disease Intervention Trial). Circ. Cardiovasc. Genet. 10, (2017).

48. Wu, J. et al. A summary of the effects of antihypertensive medications on measured blood pressure. Am. J. Hypertens. 18, 935–942 (2005).

49. Esposito, K., Chiodini, P., Bellastella, G., Maiorino, M. I. & Giugliano, D. Proportion of patients at HbA1c target <7% with eight classes of antidiabetic drugs in type 2 diabetes: systematic review of 218 randomized controlled trials with 78 945 patients. Diabetes Obes. Metab. 14, 228–233 (2012).

50. Bulik-Sullivan, B. K. et al. LD Score regression distinguishes confounding from polygenicity in genome-wide association studies. Nat. Genet. 47, 291–295 (2015).

51. Friedman, J., Hastie, T. & Tibshirani, R. Regularization paths for generalized linear models via coordinate descent. J. Stat. Softw. 33, 1–22 (2010).

52. Tay, J. K., Narasimhan, B. & Hastie, T. Elastic net regularization paths for all generalized linear models. J. Stat. Softw. 106, 1–31 (2023).

