## Supplementary Note for "Exposure accumulation drives age-dependent disease architectures and polygenic risk scores"

August 2025

**4 Relationship between gene-by-age interaction and linear, exposure accumulation, and proportional amplification models** **8**

### 1 Model overview

#### 1.1 Quantitative traits

We study the following models:

$$L_{linear} = G + E + age, \tag{1}$$

$$L_{EA} = G + E + age + \sum_{a=0}^{age} E_a, \tag{2}$$

$$L_{PA} = G + E + age + age * (G + E), \tag{3}$$

where $L$ is disease liability (or quantitative trait), $G$ is the genetic component, $E$ is the environmental component, $age$ is the age quintile. EA denotes exposure accumulation and PA denotes proportional amplification.

We are interested in the liability heritability and the accuracy of using PRS to predict $L$ (measured by $R^2_{PRS}$):

$$R^2_{PRS} \propto h^2 = \rho(G, L)^2 \tag{4}$$

Covariance between G and E has an impact on $R^2_{PRS}$ and $h_2$, which we discuss in Section 2.3.

#### 1.2 Diseases

Diseases are defined as:

$$D = \begin{cases} 1, \ L_{linear} > \mathcal{T} \\ 0, \ L_{linear} \leq \mathcal{T} \end{cases} \quad (5)$$

Prediction accuracy of diseases can be measured by the square of point-biserial correlation ($R^2_{bp}$, equation 7) or liability-scale correlation $R^2_{liab}$. On empirical data, we used the observed-scale $R^2_{obs}$, which is a special case of $R^2_{bp}$ measured in samples of fixed case-control ratio. $R^2_{obs}$ is proportional to $R^2_{liab}$ when population prevalence is fixed and liability is a standard normal distribution [1].

#### 1.3 Prevalent case association and incident case prediction

We distinguish two scenarios of case-control studies under the liability threshold model. We assume all individuals are followed (i.e. without censoring) to simplify the discussion while random censoring doesn't impact our results:

- **Prevalent case association**: cases are defined as $L(age) > \mathcal{T}$, controls are defined as $L(age) \leq \mathcal{T}$. Specifically, at 60 years of age, cases are all individuals who have been diagnosed with the disease before 60, while controls are all those who were not diagnosed with the disease at 60 years of age. Prevalent case association are applied in majority of GWAS and PRS studies in statistical genetics.
- **Incident case prediction**: cases are defined as $L(age_{begin}) < \mathcal{T} \leq L(age_{end})$, controls are defined as $L(age_{end}) \leq \mathcal{T}$. Specifically, for predicting 10-year risk at 50 years of age, cases are all individuals who have got disease between 50 and 60 years of age, while controls are all those who remained healthy at 60 years of age. Incident case prediction are widely applied in clinical risk prediction and is informative for clinical decision making. In practice, individuals are censored before $age_{end}$, which needs additional treatment such as inverse-probability weighting [2] or time-to-event regression (Cox model).

Under the definition of the two case-control studies, the controls are defined in the same way, while the cases are defined differently. Age-specific prediction accuracy are usually lower for incident case prediction compared to prevalence case association, for the same age point, which we will explain in the next section (Section 2).

### 2 Description of mechanisms that lead to decreased PRS prediction accuracy with age

We found three mechanisms that can lead to decreasing PRS prediction accuracy in simulations and analyses of empirical data, which can be summarised by following three inequalities:

- $h^2_{young} = cor(G_{young}, L_{young})^2 > h^2_{old} = cor(G_{old}, L_{old})^2$.
- $E[G_{case,young}] - E[G_{control,young}] > E[G_{case,old}] - E[G_{control,old}]$.
- $R^2_{liab,incident}(Young) > R^2_{liab,incident}(Old)$.

The first mechanism is trivial mathematically. Section 3 provides proofs of the second and third mechanisms. We note there is **mechanism 4** which is the overall prediction accuracy irrespective of age ($R^2_{liab}$) can cause different age dependency. We discuss this mechanism separately after equation 7 as it does not cause **decreases** of PRS prediction accuracy.

#### 2.1 Mechanism 1: Liability-scale heritability changes with age

Under this mechanism, the liability-scale heritability ($h^2_{liab}$) changes with age through changes in environmental variance or the genetic variance. $h^2_{liab}$ can decrease when the genetic variance decreases or when the environmental variance increases (The EA model). PRS prediction accuracy decreases with $h^2_{liab}$ (equation 4). We note this mechanism can only be tested in simulations for disease traits, as we can not estimate $h^2_{liab}$ for age interval as the assumptions are based in the entire population [1].

#### 2.2 Mechanism 2: liability of older cases is closer to healthy controls than younger cases

Under the liability-threshold model, a individual who get disease at 75 years of age is much healthier than an individual who get disease at 40 years of age. This intuition can be formally expressed as the mean of liability or genetic value of older cases are closer to healthy controls, which makes it harder for models to distinguish them.

#### 2.3 Mechanism 3: Collider bias due to removal of past prevalent cases

Under **incident case prediction**, individuals who have get the disease before baseline are not included in the prediction of future cases. This creates a collider bias on the liability scale, which induces a negative correlation between $G$ and $E$. We derive the expected reduction in $R^2_{liab}$ under this model below.

We define a collider, $C$, which takes a value of 1 when an individual has prevalent disease and 0 when they do not. The correlation between $G$ and $E$ stratified by $C$ can be expressed as: $cov(G, E|C = 0) = E[GE|C = 0] - E[G|C = 0] * E[E|C = 0]$. Without the collider, $\text{cov}(G, E) = 0$. Prediction accuracy of PRS is expressed as:

$$\begin{aligned} R_{liab} \propto \sqrt{h^2_{liab}} = \rho(G, L) &= \frac{V(G) + \text{cov}(G, E)}{\sqrt{(V(G) + V(E) + 2\text{cov}(G, E))V(G)}} \\ &= \frac{V(G) + \text{cov}(G, E)}{\sqrt{V(L)V(G)}} \end{aligned} \tag{6}$$

The PRS accuracy is expected to decrease because of the negative correlation between $G$ and $E$, which we will prove in section 3.3.

### 3 Math of age-dependent prediction

We group most proofs in this section. Section 3.1 discusses expression of three important metrics used in the paper; Section 3.2 presents mathematical properties of liability threshold model with $G$ and $E$ and proof of Theorem 1 that establishes decreasing prediction accuracy of mechanism 2.2; Section 3.3 presents the analytical expression of covariance between $G$ and $E$ in incident case prediction.

#### 3.1 Liability-scale $R^2$, AUC, and observed-scale $R^2$

We measured the age-specific prediction accuracy using three metrics ($R^2_{liab}$, $AUC$, and $R^2_{obs}$) that are suitable under the liability-threshold model. In practice, we found age-dependent prediction accuracy has similar pattern when measured by log odds ratio and Cox hazard ratio.

**Liability-scale** $R^2$ ($R^2_{liab}$) is defined as the correlation between liability and a predictor. When using $\hat{G}$ to represent PRS, $R^2_{liab} = \rho(\hat{G}, L)^2 = \left(\sqrt{var(\hat{G})} + cov(\hat{G}, E)/\sqrt{var(\hat{G})}\right)^2$. For prevalent case association under the Linear or $Linear_{LT}$ models, $R^2_{liab}$ does not depend on $age$. For incident disease prediction, $cov(\hat{G}, E) = cov(\hat{G}, E|L(age_{begin}) \le \mathcal{T}) < 0$ (see derivation in section 3.3).

**AUC** is defined as the probability that a randomly sampled case has a score that are higher than that of a randomly sampled control $AUC = \mathbf{P}(G_{case} > G_{control})$. We can approximate the distribution of $G_{case}$ and $G_{control}$ using Laplace approximation (normal approximation) and get an approximated AUC:

$$AUC(a) \approx \Phi(\frac{E[G|L > a] - E[G|L \le a])}{\sqrt{var(E[G|L > a]) + var(E[G|L \le a])}})$$

$\Phi$ is the CDF of standard normal distribution. Numerator within $\Phi$ strictly decrease with $a$ (**Theorem 1**); this provides the intuition that $AUC$ decreases with age under this model. Numeric simulation can establish the monotonic decrease of AUC by sampling scores of cases and controls using importance sampling of based on equation 8 ($f_{X_1|L>a}(x)$); note we need to substitute $G$ into $X_1$ in equation 8.

**Observed-scale** $R^2$ is the square of the point-biserial correlation coefficient $R_{bp}$:

$$\begin{aligned} R_{bp} &= \frac{cov(\mathbf{1}_{L>a}, G)}{\sqrt{var(\mathbf{1}_{L>a}), var(G)}} \\ &= \frac{E\left[\mathbf{1}_{L>a} G\right] - E\left[\mathbf{1}_{L>a}\right] E\left[G\right]}{\sqrt{var(\mathbf{1}_{L>a}), var(G)}} \\ &= \frac{E\left[G|L>a\right] p - p\Big(pE\left[G|L>a\right] + (1-p)E\left[G|L\leq a\right]\Big)}{\sqrt{var(\mathbf{1}_{L>a}), var(G)}} \\ &= \frac{p\left(1-p\right)\left(E\left[G|L>a\right] - E\left[G|L\leq a\right]\right)}{\sqrt{var(\mathbf{1}_{L>a}), var(G)}} \\ &= \frac{\sqrt{p\left(1-p\right)}}{\sqrt{var(G)}}\left(E\left[G|L>a\right] - E\left[G|L\leq a\right]\right), \end{aligned} \tag{7}$$

where $p = E\left[\mathbf{1}_{L>a}\right]$ is the prevalence of the disease in the target population. $R_{bp}$ is proportional to the difference between the expectation of $G$ in cases and controls ($E\left[G|L>a\right] - E\left[G|L\leq a\right]$). The derivative of ($E\left[G|L>a\right] - E\left[G|L\leq a\right]$) w.r.t $a$ is strictly positive when $a > \mu_1 + \mu_2$ (Theorem 1), which means $R^2_{bp}$ decreases with age increases when $\frac{\sqrt{(p(1-p)}}{\sqrt{var(G)}}$ is constant across age. Relating to **Mechanism 4**, for $G$ with different liability-scale $R^2_{liab}(G)$, $var(G)$ will have different age-dependency. High $R^2_{liab}(G)$ will results in $var(G)$ changing with $\Delta(a)$ (defined below), which reduce the age-dependency. Intuitively, predictors with low $R^2_{liab}(G)$ will have similar $var(G)$ at different age point as most variation in $G$ is not related to disease status. Age-dependent $var(G)$ can be verified using numeric simulation, while we skip its analytical derivation.

#### 3.2 Bivariate normal distribution of $X_1$ and $X_2$

We first repeated the derivation of several well-known quantities of bivariate normal distribution:

$$\begin{pmatrix} X_1 \\ X_2 \end{pmatrix} \sim N\left(\begin{pmatrix} \mu_1 \\ \mu_2 \end{pmatrix}, \begin{pmatrix} \sigma_1^2 & 0 \\ 0 & \sigma_2^2 \end{pmatrix}\right).$$

First, PDF of $L = X_1 + X_2$ is:

$$f_L(l) = \int_{-\infty}^{\infty} f_{X_1}(x) f_{X_2}(l-x)dx = \frac{1}{\sqrt{2\pi(\sigma_1^2+\sigma_2^2)}} \exp\left(-\frac{(l-(\mu_1+\mu_2))^2}{2((\sigma_1^2+\sigma_2^2))}\right).$$

The above results are derived by simplifying terms within exp to extract the quadratic terms of $x$, then gathering remaining terms w.r.t. $l$ (which will give a quadratic term of $l$). Using the fact that $f_L(l)$ is a probability distribution w.r.t. exponential of a quadratic term of $l$, $f_L(l)$ is a normal distribution. In fact, this could be derived from a more general results of affine transformation of normal distribution, see equation 2.113-2.117 of [3].

As a corollary, we derive the truncated probability of $L$:

$$\mathbf{P}(L < a) = \Phi(\frac{(a - (\mu_1 + \mu_2))^2}{\sqrt{\sigma_1^2 + \sigma_2^2}}),$$

where $\Phi$ is the CDF of standard normal distribution.

Next, we derive $f(X_1|L \leq a)$ and $f(X_1|L > a)$, which is straightforward rescaling of the truncated distribution:

$$\begin{aligned} f_{X_1|L\leq a}(x) &= \frac{f_{X_1}(x)\Phi((a-x-\mu_2)/\sigma_2)}{\mathbf{P}(L<a)} \\ f_{X_1|L>a}(x) &= \frac{f_{X_1}(x)\left(1-\Phi((a-x-\mu_2)/\sigma_2)\right)}{1-\mathbf{P}(L<a)} \end{aligned} \tag{8}$$

In previous sections, we repeated use the difference of genetic values between cases and controls. Therefore, in the following part we focus on deriving the expectation $E[X_1|L < a]$, expectation $E[X_1|L \geq$

$a$], and the difference $\Delta(a) = E[X_1|L \geq a] - E[X_1|L < a]$. First we notice vector $(L, X_1)$ is a linear transformation of $(X_1, X_2)$, therefore it is also a joint normal distribution. Following the distribution of affine mapping of normal distribution, we note the joint distribution of $(L, X_1)$ is:

$$\begin{pmatrix} X_1 \\ X_1 + X_2 \end{pmatrix} \sim N\left(A\begin{pmatrix}\mu_1 \\ \mu_2\end{pmatrix}, A\begin{pmatrix}\sigma_1^2 & 0 \\ 0 & \sigma_2^2\end{pmatrix}A^T\right) = N\left(\begin{pmatrix}\mu_1 \\ \mu_1 + \mu_2\end{pmatrix}, \begin{pmatrix}\sigma_1^2 & \sigma_1^2 \\ \sigma_1^2 & \sigma_1^2 + \sigma_2^2\end{pmatrix}\right). \tag{9}$$

Here we use $A = \begin{pmatrix}1 & 0 \\ 1 & 1\end{pmatrix}$. Using equation 2.79, 2.80, 2.96, and 2.97 of [3], the conditional distribution of $X_1|X_1 + X_2 = y$ is a normal distribution:

$$\begin{aligned} P(X_1|X_1 + X_2 = y) &= \mathcal{N}(\mu_1 + \frac{\sigma_1^2}{\sigma_2^2 + \sigma_1^2}(y - (\mu_1 + \mu_2), \frac{\sigma_1^2\sigma_2^2}{\sigma_2^2 + \sigma_1^2}) \\ E[X_1|X_1 + X_2 = y] &= \mu_1 + \frac{\sigma_1^2}{\sigma_2^2 + \sigma_1^2}(y - (\mu_1 + \mu_2)) \end{aligned} \tag{10}$$

We notice $E[X_1|X_1 + X_2 = y]$ is a linear function of $y$. Therefore, using $L = X_1 + L_2$, the truncated distribution $E[X_1|L < a] = E[E[X_1|L]|L < a] = \mu_1 + \frac{\sigma_1^2}{\sigma_2^2+\sigma_1^2}(E[L|L < a] - (\mu_1 + \mu_2))$. We note the last term $(E[L|L < a] - (\mu_1 + \mu_2))$ can be expressed as inverse Mills ratio: $(E[L|L < a] - (\mu_1 + \mu_2)) = -\sqrt{\sigma_2^2 + \sigma_1^2}\frac{\phi(\alpha)}{\Phi(\alpha)}, \alpha = \frac{a-(\mu_1+\mu_2)}{\sqrt{\sigma_2^2+\sigma_1^2}}$. Here $\phi$ is the pdf of standard normal distribution and $\Phi$ is the CDF of standard normal distribution. Therefore, the expression of $E[X_1|L < a]$ is:

$$E[X_1|L < a] = \mu_1 - \frac{\sigma_1^2}{\sqrt{\sigma_2^2 + \sigma_1^2}}\frac{\phi(\alpha)}{\Phi(\alpha)}.$$

Similarly, since $(E[L|L \geq a] - (\mu_1 + \mu_2)) = \sqrt{\sigma_2^2 + \sigma_1^2}\frac{\phi(\alpha)}{1-\Phi(\alpha)}, \alpha = \frac{a-(\mu_1+\mu_2)}{\sqrt{\sigma_2^2+\sigma_1^2}}$, we have:

$$E[X_1|L \geq a] = \mu_1 + \frac{\sigma_1^2}{\sqrt{\sigma_2^2 + \sigma_1^2}}\frac{\phi(\alpha)}{1 - \Phi(\alpha)}.$$

The difference between $E[X_1|L \geq a]$ and $E[X_1|L < a]$ is:

$$\begin{aligned} \Delta(a) &= E[X_1|L \geq a] - E[X_1|L < a] \\ &= \frac{\sigma_1^2}{\sqrt{\sigma_2^2 + \sigma_1^2}}\left(\frac{\phi(\alpha)}{1 - \Phi(\alpha)} + \frac{\phi(\alpha)}{\Phi(\alpha)}\right) \\ &= \frac{\sigma_1^2}{\sqrt{\sigma_2^2 + \sigma_1^2}}\left(\lambda_+(\alpha) + \lambda_-(\alpha)\right). \end{aligned} \tag{11}$$

Here we use $\lambda_+$ and $\lambda_-$ to refer to the inverse Mills ratio. We are most interested in the derivative of $\Delta(a)$ w.r.t. the threshold $a$. Noticing $\alpha = \frac{a-(\mu_1+\mu_2)}{\sqrt{\sigma_2^2+\sigma_1^2}}$, we use the derivative w.r.t. $\alpha$ to simplify the expressions and note the sign of derivatives should be the same. Making use of the derivatives $\frac{\partial}{\partial\alpha}\lambda_+ = -\alpha\lambda_+ + (\lambda_+)^2$ and $\frac{\partial}{\partial\alpha}\lambda_- = -\alpha\lambda_- - (\lambda_-)^2$:

**Theorem 1** *For $\alpha > 0$, the sum of inverse Mills' ratios $\lambda_+ + \lambda_-$ is a monotonically increasing function:*

$$\frac{\partial}{\partial\alpha}\Delta(\alpha) = \frac{\sigma_1^2}{\sqrt{\sigma_2^2 + \sigma_1^2}}\left(\lambda_+(\alpha) + \lambda_-(\alpha)\right)\left(\lambda_+(\alpha) - \lambda_-(\alpha) - \alpha\right) > 0$$

**Proof:** Following inequalities can be obtained from Equation (9) of [4] ($\lambda_+(\alpha) > \alpha$, $\lambda_-(\alpha \to \infty) = \alpha$, $\lambda_-(\alpha \to \infty) = 0+$) and theorem 2.3 in [5]:

$$\lambda_+ - \alpha > \frac{\sqrt{\alpha^2 + 8} - \alpha}{4} > 0$$

$$\lambda_+ - \alpha < \frac{\sqrt{\alpha^2 + 4} - \alpha}{2}$$

Using $\Phi(\alpha) > 0.5$ for $\alpha > 0$:

$$\lambda_- < \sqrt{\frac{2}{\pi}} e^{-\frac{\alpha^2}{2}}.$$

The goal is showing $T(\alpha) = \lambda_+(\alpha) - \lambda_-(\alpha) - \alpha$ is positive:

$$\begin{aligned} T(0) &= 0 \\ T(\alpha \to \infty) &= 0 \\ \frac{\partial}{\partial \alpha} T(\alpha) &= -\alpha\lambda_+ + (\lambda_+)^2 + \\ & \alpha\lambda_- + (\lambda_-)^2 \\ & -1 \end{aligned} \tag{12}$$

First, we compare the lower bound of $\lambda_+ - \alpha$ with the upper bound of $\lambda_-$ for large $\alpha$ using L'Hôpital's rule:

$$\lim_{\alpha \to \infty} \frac{(\frac{\sqrt{\alpha^2+8}-\alpha}{4})'}{(\sqrt{\frac{2}{\pi}} e^{-\frac{\alpha^2}{2}})'} = \frac{0.25 \times (\frac{\alpha}{\sqrt{\alpha^2+8}} - 1)}{\sqrt{\frac{2}{\pi}} \times -\alpha \times e^{-\frac{\alpha^2}{2}}} = \infty.$$

The upper bound of $\lambda_-$ decreases more sharply than the lower bound of $\lambda_+ - \alpha$ for large $\alpha$; therefore, the lower bound of $\lambda_+ - \alpha$ is larger than the upper bound of $\lambda_-$ for large $\alpha$, suggesting $T(\alpha \to \infty) = 0+$. By plugging in numbers, we find $\frac{(\frac{\sqrt{\alpha^2+8}-\alpha}{4})'}{(\sqrt{\frac{2}{\pi}} e^{-\frac{\alpha^2}{2}})'}|_{alpha=2.6} = 1.144 > 1$ and $T(2.6) = 0.30 > 0$. Using $f$ to represent the lower bound of $\lambda_+ - \alpha$ and $g$ to represent the upper bound of $\lambda_-$, $\frac{f(a)}{g(a)} = \frac{f(a)-f(\infty)}{g(a)-g(\infty)} \approx \frac{f'(a_0 \in [a,\infty])}{g'(a_0 \in [a,\infty])}$. $f'$ dominate $g'$ for $\alpha > 2.6$ proves $T(\alpha) > 0$ for $\alpha > 2.6$.

Second, for $\alpha \in (0, 2.6)$, $\frac{\partial}{\partial \alpha} T(\alpha)|_{\alpha=0} > 0$ implies $T(\alpha)$ increases first. Next, we use numeric evaluation and found $\frac{\partial}{\partial \alpha} T(\alpha)$ crosses zero around 1.994. Inverse Mills' ratio is strictly convex on $\mathcal{R}$ [6], which implies $\frac{\partial^2}{\partial^2 \alpha}\lambda_+(\alpha) > 0$ and $\frac{\partial^2}{\partial^2 \alpha}\lambda_-(\alpha) = \frac{\partial^2}{\partial^2 \alpha}\lambda_+(-\alpha) > 0$. We can separate $T(\alpha)$ into two monotonic parts:

$$\frac{\partial}{\partial \alpha}\lambda_+(\alpha) = -\alpha\lambda_+ + (\lambda_+)^2$$

$$-\frac{\partial}{\partial \alpha}\lambda_-(\alpha) = \alpha\lambda_- + (\lambda_-)^2$$

Between $(0, 1]$, $-\alpha\lambda_+ + (\lambda_+)^2$ increases from 0.637 to 0.801 while $\alpha\lambda_- + (\lambda_-)^2$ decreases from 0.637 to 0.370+; therefore $\frac{\partial}{\partial \alpha} T(\alpha)$ is strictly positive on $(0, 1]$ with $T(1) = 0.237+$. We repeat the same procedure between $(1, 1.6]$ and found $-\alpha\lambda_+ + (\lambda_+)^2$ increases from 0.801 to 0.858+ while $\alpha\lambda_- + (\lambda_-)^2$ decreases from 0.370 to 0.201+, again suggesting $\frac{\partial}{\partial \alpha} T(\alpha)$ is strictly positive on $(0, 1.6]$ with $T(1.6) = 0.306+$. Lastly on $(1.6, 2.6]$ we notice $\alpha\lambda_- + (\lambda_-)^2$ decreases from 0.201534 to 0.035+, which means the fluctuation of $T(\alpha)$ between $(1.6, 2.6]$ is bounded by $|T(x) - T(1.6)|_{x \in (1.6, 2.6]} < 0.17 \times (2.6 - 1.6) = 0.17 < T(1.6) = 0.306+$; this implies that $T(\alpha)$ is strictly positive on $(0, 2.6]$. Therefore we conclude the proof for $\alpha \in (0, \infty]$.

**Theorem 1** is proved for standard normal distribution but we note these results are generalisable to some other distributions [7].

**Corollary 1.1** *Under the $Linear_{LT}$ model with normal liability distribution, observed-scale R-sqaured ($R^2_{bp}$) in samples with the same genetic variance and case-control ratio strictly decreases with age.*

**Proof** Equation 7 shows $R^2_{bp}$ is a linear function of $\Delta(a)^2$ and $\frac{\sqrt{(p(1-p)}}{\sqrt{var(G)}}$. Since $a$ decreases with age, $R^2_{bp}$ also decreases with age. Corollary 1.1 suggest observed-scale $R^2_{obs}$ are comparable only when sample has fixed case-control ratio.

#### 3.3 Collider effect estimation

We use a liability-threshold model with liability-scale heritability $h^2$ and disease prevelance $\mathcal{P}$. Liability threshold assumes $L = G + E, var(G)/var(L) = h^2$, where $G$ and $E$ are independent at birth following the definition of non-genetic variance. For incident case prediction, removing prevalent cases at baseline would create negative correlation between $G$ and $E$.

Using $C$ to denote the prevalent diseases $C = 1 : L = G + E > \mathcal{T}$, where $\mathcal{T}$ is the threshold above which an individual become a case. We are interested in the covariance of $G$ and $E$ conditional on not having the disease at baseline:

$$\mathrm{cov}(G, E|C = 0) = E[G \cdot E|C = 0] - E[G|C = 0]E[E|C = 0]. \tag{13}$$

The derivation of $E[G \cdot E|C = 0]$ uses the integration of truncated bivariate Gaussian distribution, which we derived below as preparation:

$$\begin{aligned}
A &= \int_{-\infty}^{\infty}\int_{-\infty}^{\mathcal{T}-x_1} x_1 x_2 N(x_1|0,\sigma_1^2)N(x_2|0,\sigma_2^2)dx_1dx_2 \\
&= \int_{-\infty}^{\infty} x_1 \left[\int_{-\infty}^{\mathcal{T}-x_1} x_2 N(x_2|0,\sigma_2^2)dx_2\right] N(x_1|0,\sigma_1^2)dx_1, \\
A_1 &= \left[\int_{-\infty}^{\mathcal{T}-x_1} x_2 N(x_2|0,\sigma_2^2)dx_2\right] \\
&= (-\sqrt{\frac{\sigma_2^2}{2\pi}})\int_{-\infty}^{\mathcal{T}-x_1} d\left(\exp(-\frac{x_1^2}{2\sigma_2^2})\right) \\
&= (-\sqrt{\frac{\sigma_2^2}{2\pi}})\left(\exp(-\frac{(x_1-\mathcal{T})^2}{2\sigma_2^2})\right), \\
A &= (-\sqrt{\frac{\sigma_2^2}{2\pi}})\int_{-\infty}^{\infty} x_1 N(x_1|0,\sigma_1^2)\exp(-\frac{(x_1-\mathcal{T})^2}{2\sigma_2^2})dx_1 \qquad (14)\\
&= (-\sqrt{\frac{\sigma_2^2}{2\pi}}\cdot\sqrt{\frac{1}{2\pi\sigma_1^2}})\int_{-\infty}^{\infty} x_1\exp\left(-\frac{x_1}{2\sigma_1^2}-\frac{(x_1^2-\mathcal{T})^2}{2\sigma_2^2}\right)dx_1 \\
&= (-\sqrt{\frac{\sigma_2^2}{2\pi}}\cdot\sqrt{\frac{1}{2\pi\sigma_1^2}})\int_{-\infty}^{\infty} x_1\exp\left(-\frac{1}{2}(\frac{1}{\sigma_1^2}+\frac{1}{\sigma_2^2})(x_1-\mathcal{T}\frac{\sigma_1^2}{\sigma_1^2+\sigma_2^2})^2-\frac{\mathcal{T}^2}{2\sigma_2^2}+\frac{\mathcal{T}^2}{2\sigma_2^2}\frac{\sigma_1^2}{\sigma_1^2+\sigma_2^2}\right)dx_1 \\
&= (-\sqrt{\frac{\sigma_2^2}{2\pi}}\cdot\sqrt{\frac{1}{2\pi\sigma_1^2}}\cdot\exp\left(-\frac{\mathcal{T}^2}{2(\sigma_1^2+\sigma_2^2)}\right))\int_{-\infty}^{\infty} x_1\exp\left(-\frac{1}{2}(\frac{1}{\sigma_1^2}+\frac{1}{\sigma_2^2})(x_1-\mathcal{T}\frac{\sigma_1^2}{\sigma_1^2+\sigma_2^2})^2\right)dx_1 \\
&= -\sqrt{\frac{\sigma_2^2}{2\pi}}\cdot\sqrt{\frac{1}{2\pi\sigma_1^2}}\cdot\exp\left(-\frac{\mathcal{T}^2}{2(\sigma_1^2+\sigma_2^2)}\right)\cdot\sqrt{2\pi\frac{\sigma_1^2\sigma_2^2}{\sigma_1^2+\sigma_2^2}}\mathcal{T}\frac{\sigma_1^2}{\sigma_1^2+\sigma_2^2} \\
&= -\sqrt{\frac{1}{2\pi}}\cdot\mathcal{T}\frac{\sigma_1^2\sigma_2^2}{(\sigma_1^2+\sigma_2^2)^{\frac{3}{2}}}\cdot\exp\left(-\frac{\mathcal{T}^2}{2(\sigma_1^2+\sigma_2^2)}\right).
\end{aligned}$$

Similarly, we derive:

$$\begin{aligned}
B &= \int_{-\infty}^{\infty}\int_{-\infty}^{\mathcal{T}-x_1} x_2 N(x_1|0,\sigma_1^2)N(x_2|0,\sigma_2^2)dx_1dx_2 \\
&= (-\sqrt{\frac{\sigma_2^2}{2\pi}}\cdot\sqrt{\frac{1}{2\pi\sigma_1^2}}\cdot\exp\left(-\frac{\mathcal{T}^2}{2(\sigma_1^2+\sigma_2^2)}\right))\int_{-\infty}^{\infty}\exp\left(-\frac{1}{2}(\frac{1}{\sigma_1^2}+\frac{1}{\sigma_2^2})(x_1-\mathcal{T}\frac{\sigma_1^2}{\sigma_1^2+\sigma_2^2})^2\right)dx_1 \qquad (15)\\
&= -\frac{\sigma_2^2}{\sqrt{2\pi(\sigma_1^2+\sigma_2^2)}}\cdot\exp\left(-\frac{\mathcal{T}^2}{2(\sigma_1^2+\sigma_2^2)}\right).
\end{aligned}$$

**Theorem 2** *Under liability threshold model, the population without prevalent cases has negative (liability-scale) correlations of $G$ and $E$. The covariance has following relationship to liability-scale heritability and disease prevalence* $= 1 - \Phi(\mathcal{T})$

$$cov(G, E|C = 0) = -\frac{h^2(1-h^2)}{\sqrt{2\pi}}\cdot\exp\left(-\frac{\mathcal{T}^2}{2}\right)\cdot\left[\mathcal{T}+\sqrt{\frac{1}{2\pi}}\exp\left(-\frac{\mathcal{T}^2}{2}\right)\right]$$

**Proof:** Plugging above derivation of $A$ and $B$ to equation 13:

$$\begin{aligned}\text{cov}(G,E|C=0) &= -\sqrt{\frac{1}{2\pi}\frac{\sigma_1^2\sigma_2^2}{\sigma_1^2+\sigma_2^2)}}\cdot\exp\left(-\frac{\mathcal{T}^2}{2(\sigma_1^2+\sigma_2^2)}\right)\cdot\left[\frac{\mathcal{T}}{\sqrt{\sigma_1^2+\sigma_2^2}}+\sqrt{\frac{1}{2\pi}}\exp\left(-\frac{\mathcal{T}^2}{2(\sigma_1^2+\sigma_2^2)}\right)\right]\\ &= -\frac{h^2(1-h^2)}{\sqrt{2\pi}}\cdot\exp\left(-\frac{\mathcal{T}^2}{2}\right)\cdot\left[\mathcal{T}+\sqrt{\frac{1}{2\pi}}\exp\left(-\frac{\mathcal{T}^2}{2}\right)\right].\end{aligned} \tag{16}$$

Here we plugged in $\sigma_1^2 = h^2$ and $\sigma_2^2 = 1-h^2$ and uses expression of $A$ to compute $E[G\cdot E|C=0]$ and $B$ to compute $E[G|C=0]$ and $E[E|C=0]$.

**Corollary 2.1** *Under the Linear liability-threshold model with normal liability distribution, the same predictor has higher $R^2_{liab}$ for prevalent case association than incident case prediction.*

The incident case prediction effect of G on disease liability can be expressed as $\beta \propto \rho(G,L|C=0) = \sqrt{var(G)} + cov(G,E|C=0)/\sqrt{var(G)}$. Therefore, the relative decrease of association effect size is $-\frac{(1-h^2)}{\sqrt{2\pi}}\cdot\exp\left(-\frac{\mathcal{T}^2}{2}\right)\cdot\left[\mathcal{T}+\sqrt{\frac{1}{2\pi}}\exp\left(-\frac{\mathcal{T}^2}{2}\right)\right]$.

### 4 Relationship between gene-by-age interaction and linear, exposure accumulation, and proportional amplification models

Gene-age interactions (GxAge) usually defined as $Y = G + age + \beta_{G\times age}G\times age$ [8, 9]. The existence of GxAge is defined by $\beta_{G\times age} \neq 0$, which has following relationship to the three models we studied:

1. Linear model = no GxAge.
2. Exposure accumulation (EA) model = no GxAge
3. Proportional amplification (PA) model = GxAge. When applying a random effect model, such as GCI-GREML [8] to estimate the interaction component, the PA model will cause a positive GxAge signal. In detail, we can derive the expression of $V$ on the top of page 77 in Yang 2011 AJHG [10] under the PA model:
   $V(Y_{young}, Y_{young}) = cov(L_{young}, L_{young}) = \sqrt{var(L_{young})\times var(L_{young})}cor(L_{young}, L_{young})$ while $V(Y_{young}, Y_{old}) = cov(L_{young}, L_{old}) = \sqrt{var(L_{young})\times var(L_{old})}cor(L_{young}, L_{old})$; $L_{young}$ and $L_{old}$ is the random liability for young or old population.
   Under PA, $cor(L_{young}, L_{young}) = cor(L_{young}, L_{old}) = 1$ and $var(L_{young}) \neq var(L_{old})$, which means the interaction term ($g_{gci}$ in [8]) will capture additional variance.

### References

[1] Sang Hong Lee et al. "A better coefficient of determination for genetic profile analysis". In: *Genetic epidemiology* 36.3 (2012), pp. 214–224.

[2] David M Vock et al. "Adapting machine learning techniques to censored time-to-event health record data: A general-purpose approach using inverse probability of censoring weighting". In: *Journal of biomedical informatics* 61 (2016), pp. 119–131.

[3] Christopher M Bishop and Nasser M Nasrabadi. *Pattern recognition and machine learning.* Vol. 4. 4. Springer, 2006.

[4] Robert D Gordon. "Values of Mills' ratio of area to bounding ordinate and of the normal probability integral for large values of the argument". In: *The Annals of Mathematical Statistics* 12.3 (1941), pp. 364–366.

[5] Árpád Baricz. "Mills' ratio: Monotonicity patterns and functional inequalities". In: *Journal of Mathematical Analysis and Applications* 340.2 (2008), pp. 1362–1370.

[6] Michael R Sampford. "Some inequalities on Mill's ratio and related functions". In: *The Annals of Mathematical Statistics* 24.1 (1953), pp. 130–132.

[7] Arpád Baricz. “Mills’ ratio: Reciprocal convexity and functional inequalities”. In: *arXiv preprint arXiv:1010.3267* (2010).

[8] Matthew R Robinson et al. “Genotype–covariate interaction effects and the heritability of adult body mass index”. In: *Nature genetics* 49.8 (2017), pp. 1174–1181.

[9] Jiacheng Miao et al. “PIGEON: a statistical framework for estimating gene–environment interaction for polygenic traits”. In: *Nature Human Behaviour* (2025), pp. 1–15.

[10] Jian Yang et al. “GCTA: a tool for genome-wide complex trait analysis”. In: *The American Journal of Human Genetics* 88.1 (2011), pp. 76–82.
