## Supplementary Figures for "Exposure accumulation drives age-dependent disease architectures and polygenic risk scores"

**Supplementary Tables Captions**

Supplementary Table 1: Full list of models that explain the age-dependent genetic architecture of disease liability. $\Delta h^2$ is the change of heritability w.r.t. age. $\Delta var(L)$ is the change of liability variance w.r.t. age. $\Delta var(G)$ is the change of genetic variance w.r.t. age. $\rho(G_{young}, G_{old})$ is the genetic correlation between the young and old group. $\rho(E_{young}, E_{old})$ is the environmental correlation between young and old. Note we only consider a single effect in each model while complex traits can have combinations of effects (e.g. EA + PA). PA model will not impact the prediction accuracy in disease traits as it doesn't change the rank of liability in the population (Supplementary Note).

| Model | Name | GxAge | $\Delta h^2$ | $\Delta var(L)$ | $\Delta var(G)$ | $\rho(G_{young}, G_{old})$ | $\rho(E_{young}, E_{old})$ |
|---|---|---|---|---|---|---|---|
| $L = G + E + age$ | Linear | No | 0 | 0 | 0 | 1 | 1 |
| $L = G + E + age + \sum_{a=0}^{age} E_a$ | Exposure Accumulation (EA) | No | - | + | 0 | 1 | <1 |
| $L = G + E + age + age * (G + E)$ | Proportional amplification (PA) | Yes | 0 | +, - | +, - | 1 | 1 |
| $L = G_1(age) + G_2(age) + E + age$ | Age-dependent G | Yes | - | 0, +, - | 0, +, - | <1 | 1 |
| $L = G + E + age + \sum_{a=0}^{age} e^{-\gamma*(age-a)} E_a$ | Exposure accumulation with decay | No | 0, - | 0, + | 0 | 1 | <1 |
| $L = G + E + age + \beta_G * age * G + \beta_E * age * E$ | Non-proportional amplification | Yes | +, - | +,- | +, - | 1 | 1 |

Supplementary Table 2. List of 16 quantitative traits and their heritability estimates.

Supplementary Table 3. List of 9 representative diseases and their related quantitative traits. We computed both observed-scale heritability and liability-scale heritability for each disease.

Supplementary Table 4. List of 36 quantitative traits that are used to compute predicted liability of diseases.

Supplementary Table 5. Numeric values for Figure 1A. P values and Bonferroni corrected P values are reported on testing a non-zero change w.r.t age.

Supplementary Table 6. Numeric values for Figure 1B. P values and Bonferroni corrected P values are reported on testing a non-zero change w.r.t age.

Supplementary Table 7. Numeric values for Figure 1C. P values and Bonferroni corrected P values are reported for testing a non-zero change of G w.r.t age.

Supplementary Table 8. Numeric values for Figure 2A. P values are reported for testing genetic correlation different from 1 between young and old groups. Significance is determined by $p < 0.1/16$.

Supplementary Table 9. Numeric values for Figure 2B. P-values are reported for testing genetic correlation different from 1 between young and old groups. Significance is determined by Bonferroni < 0.1.

Supplementary Table 10. Accuracy of predicted disease liability. Observed-scale and liability-scale $R^2$ between predicted liability and disease status (union of both incident cases and prevalent cases) were estimated. LASSO 2-se model refers to the selection of regularisation causing the cross validation MSE to be 2 s.e. higher than the model that achieved the lowest MSE. The default setting is 1-se model while we chose 2-se as it is less likely to be affected by overfitting.

Supplementary Table 11. Numeric values for Figure 3A. For each baseline visit age quintile, we estimate heritability of predicted liability and use the same gaussian model to estimate the slopes. P-values are obtained through likelihood ratio tests. Significance was determined by $p<0.1/9$.

Supplementary Table 12. Numeric values for Figure 3B. P-values are reported for testing genetic correlation different from 1 between young and old groups for the nine disease liability. Significance is determined by $p < 0.1/9$.

Supplementary Table 13. Numeric values for Figure 4. For each disease, cases are devided into 5 quintiles based on age at diagnosis; controls are randomly sampled (excluding prevalent cases) so that case-control ratio is 1:1 for each age-quintile. We report the PRS accuracy for each age quintile using Cox Hazard Ratio, Observed-scale $R^2$, log Odds Ratio, and linear regression per-SD effect, along with their standard errors.

Supplementary Table 14. Slope of PRS ${R^2}_{obs}$ for prevalent case association vs incident case association across nine diseases. Incident cases are divided into 5 quintiles based on age at diagnosis. We use the median age of incident case age quintile as the index age to define prevalent cases as all individuals who have a diagnosis before index age. We sample controls so that case-control ratio is 1:1 for both incident case prediction and prevalent case association in each age-quintile. Slopes are estimated using a linear model with age and P-values are obtained through likelihood ratio tests. Significance was determined by $p<0.1/9$.

Supplementary Table 15. Numeric values for Figure 6A. PRS and accuracy-matched QRS has similar ${R^2}_{obs}$. P-values are reported for testing the slope being different from 0 for the nine diseases. Significance is determined by $p < 0.1/9$. Z-score are the two-tailed comparison of change in ${R^2}_{obs}$ for PRS and the accuracy-matched QRS.

Supplementary Table 16. ${R^2}_{obs}$ of PRS and accuracy-matched QRS in each age quintile for the nine diseases. S.e. are estimated from Monte-Carlo sampling of cases and controls.

Supplementary Table 17. Numeric value of Figure 6C. "Change of QRS+EA $R^2$ w.r.t. age (per 10 years) " refers the the expected change of PRS $R^2$ under the liability-threshold EA model. Last column is the z-score of the absolute difference between the estimated $R^2$ change in PRS and expected $R^2$ change from the $EA_{LT}$ model.

Supplementary Table 18. Numeric value of Figure 7. For the predictor name column, QRS, PRS, PRS + QRS, "Top 3", "Top 2" are defined in the text. Other names refer to a single predictor. We report the "change of $R^2$ w.r.t. age (per 10 years)" and "liability-scale R2" (${R^2}_{liab}$) for each predictor-disease pair.

Supplementary Table 19. Prediction ${R^2}_{obs}$ in each age quintile for all predictor-disease pairs. Across all age quintiles, we use case-control ratio 1:1 to make ${R^2}_{obs}$ comparable.

### Supplementary Figures

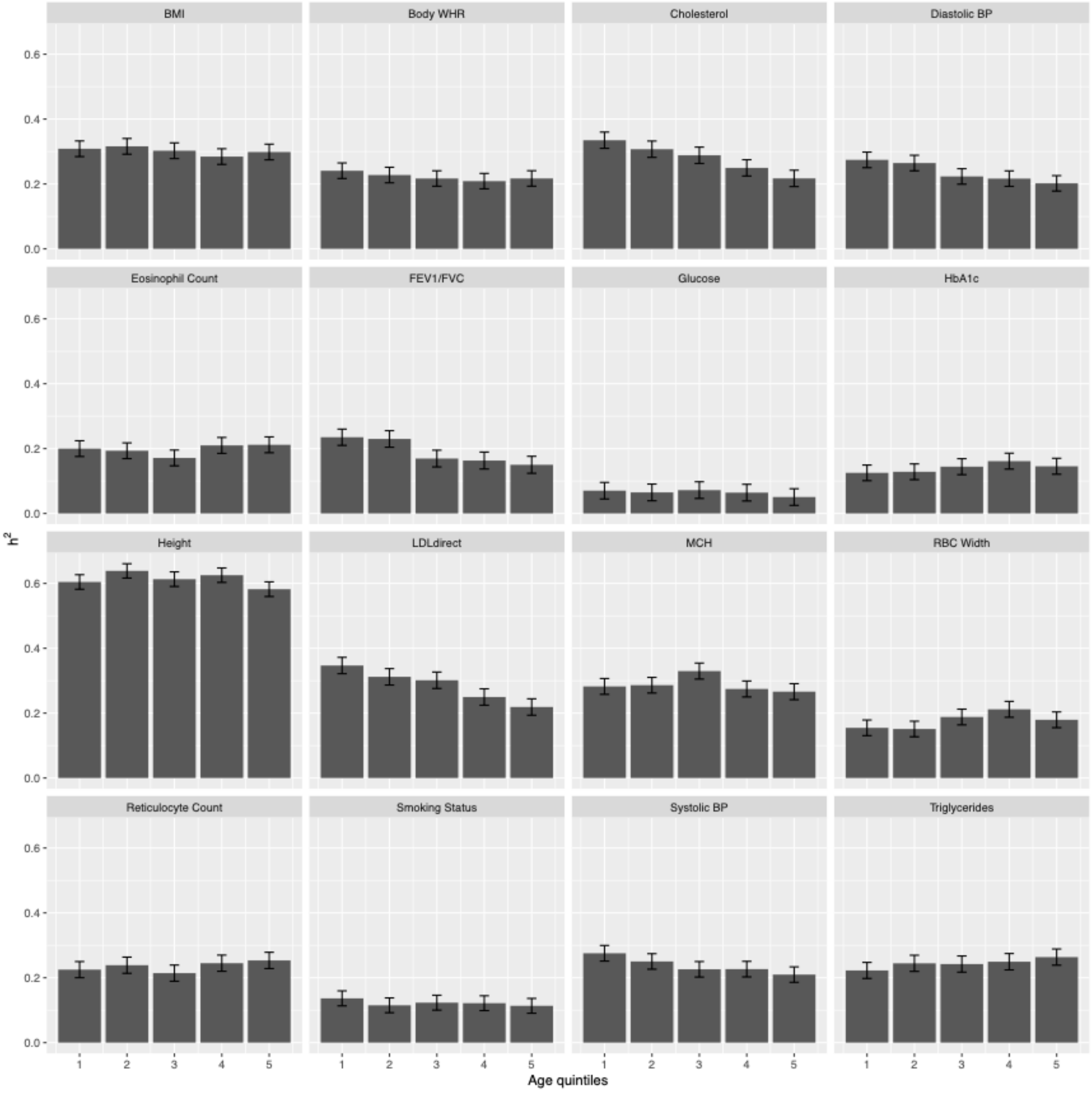


**Supplementary Figure 1. Heritability estimates of 5 age quintiles for 16 quantitative traits.** The analysis was performed using BOLT-REML, using age and genetic sex as covariates to account for within age bin age variation. Error bars are 95% confidence intervals.

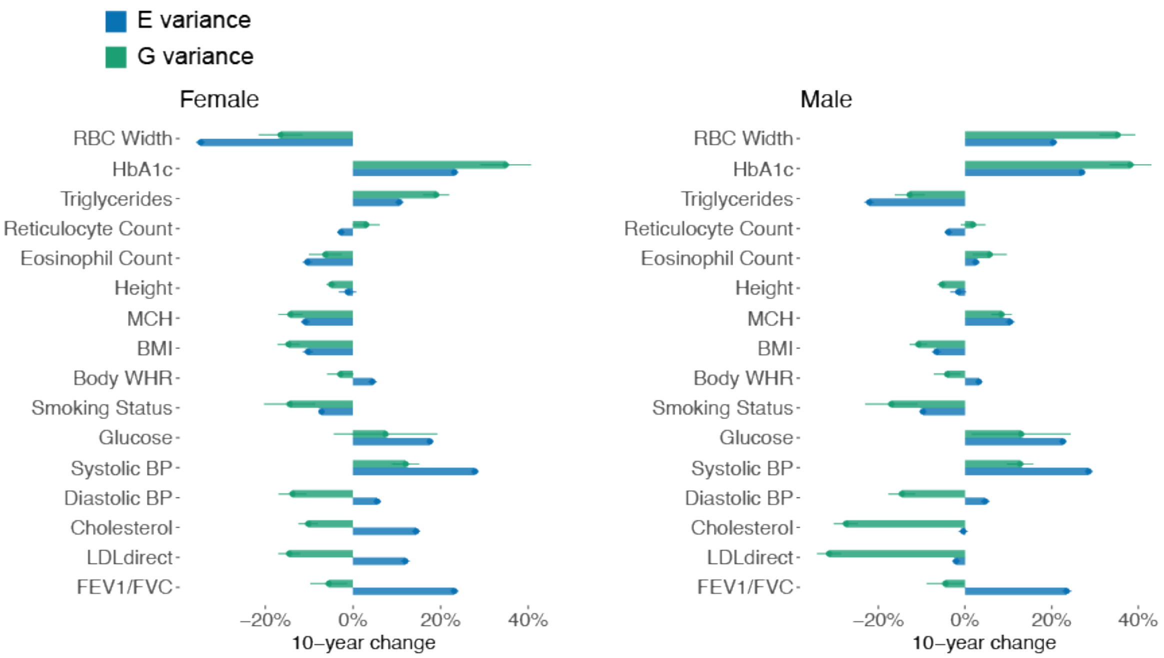


**Supplementary Figure 2: 10-year relative change for genetic (G) and environment (E) variance for the 16 quantitative traits.** Results are reported for female (left) and male (right) separately. Blue bars show the change of environment variance w.r.t age and green bars show the change of genetic variance w.r.t age. Relative changes are w.r.t. estimates in the entire population. Error bars are standard error.

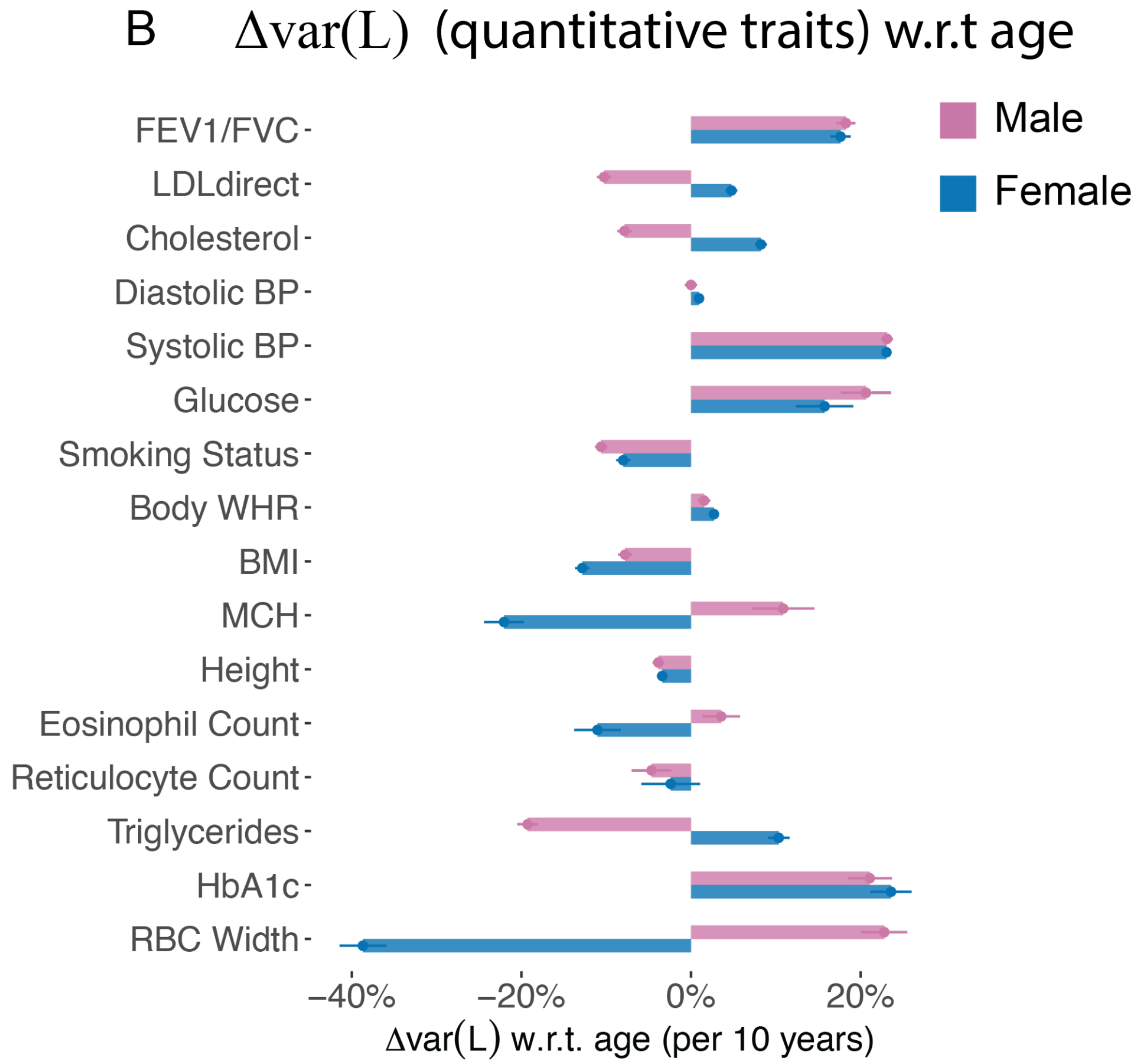


**Supplementary Figure 3.** Phenotypic variance change per 10-years for 16 quantitative traits. Purple and blue bars show female and male; error bars show standard error via bootstrapping over individuals.

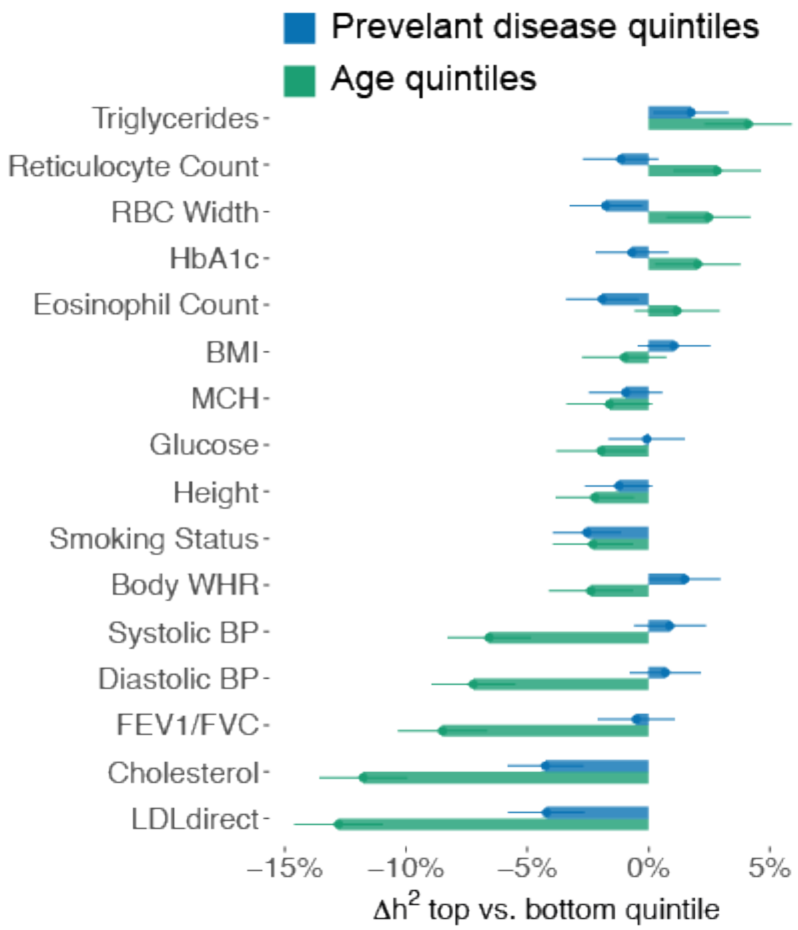


**Supplementary Figure 4. Age-dependent recruitment of healthier older people does not explain the decrease in heritability.** Error bars are standard error. Blue bars show the heritability difference between the group with highest numbers of past diseases and the group with lowest numbers of past diseases. Green bars show the heritability difference between the oldest group and the youngest group. Relative changes are w.r.t. estimates in the entire population. Error bars are standard error.

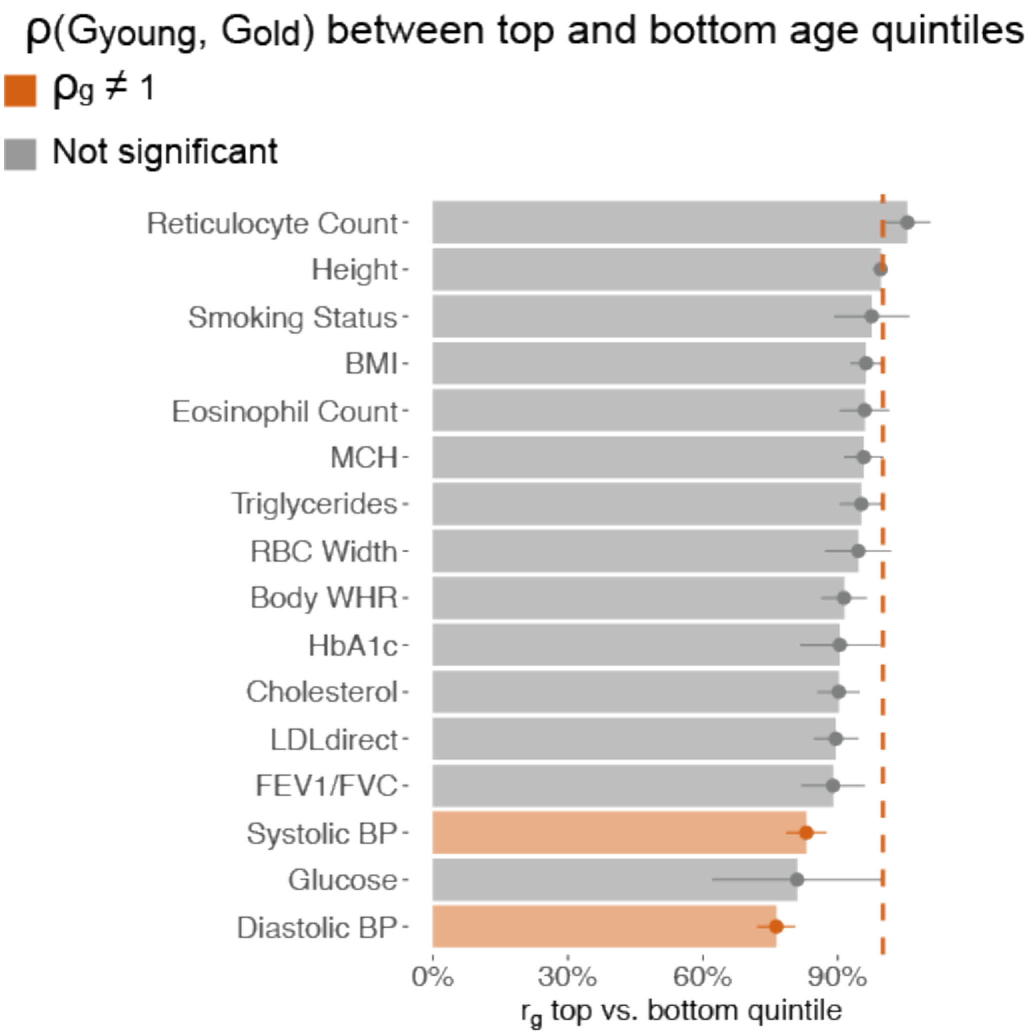


**Supplementary Figure 5. Genetic correlation between top and bottom age quintiles for 16 quantitative traits.** Traits with genetic correlation significantly different from 1 are shown in red; non-significant traits are shown in grey. Bars are standard errors. The vertical dashed line shows 1 on the x-axis.

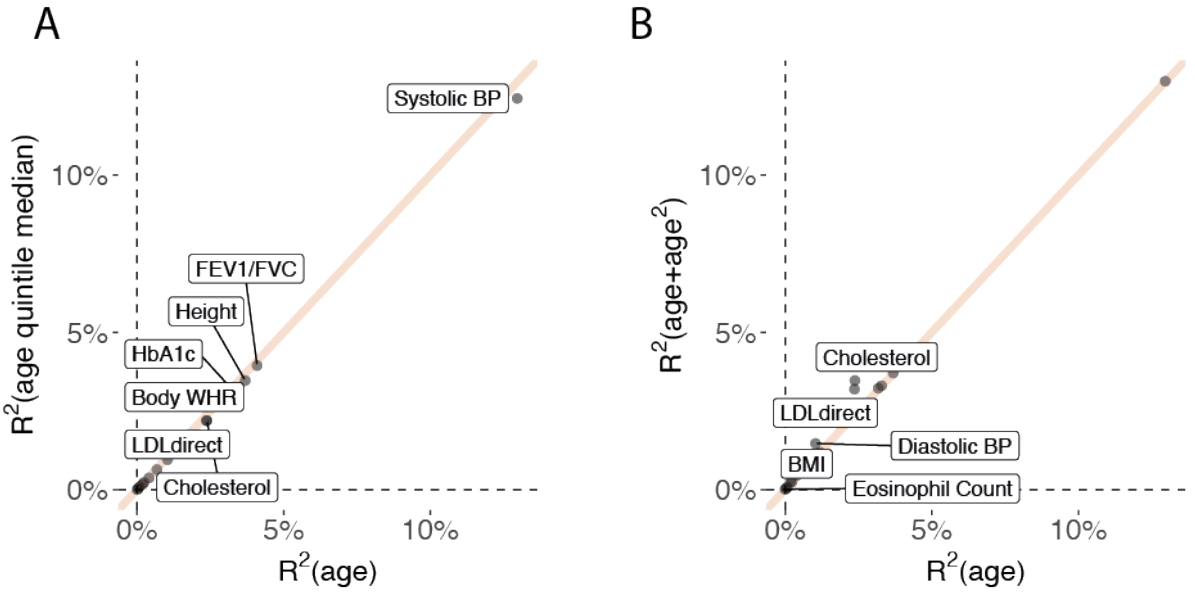


**Supplementary Figure 6. Comparison of (age in years), (age in years)², and (age quintiles) in explaining quantitative traits.** (A) We compute $R^2$ using (age in years) to predict quantitative traits and using the (median of age quintile) to predict quantitative traits. We regressed out genetic sex from each quantitative trait. Traits with prediction $R^2 > 3\%$ using (age in years) are shown for visualization. (B) We compute $R^2$ using age in years to predict quantitative traits and using (age in years) + (age in years)² to predict quantitative traits. Labeled points are those where (age in years) + (age in years)² has $R^2$ more than 1.2x than that of (age in years) only.

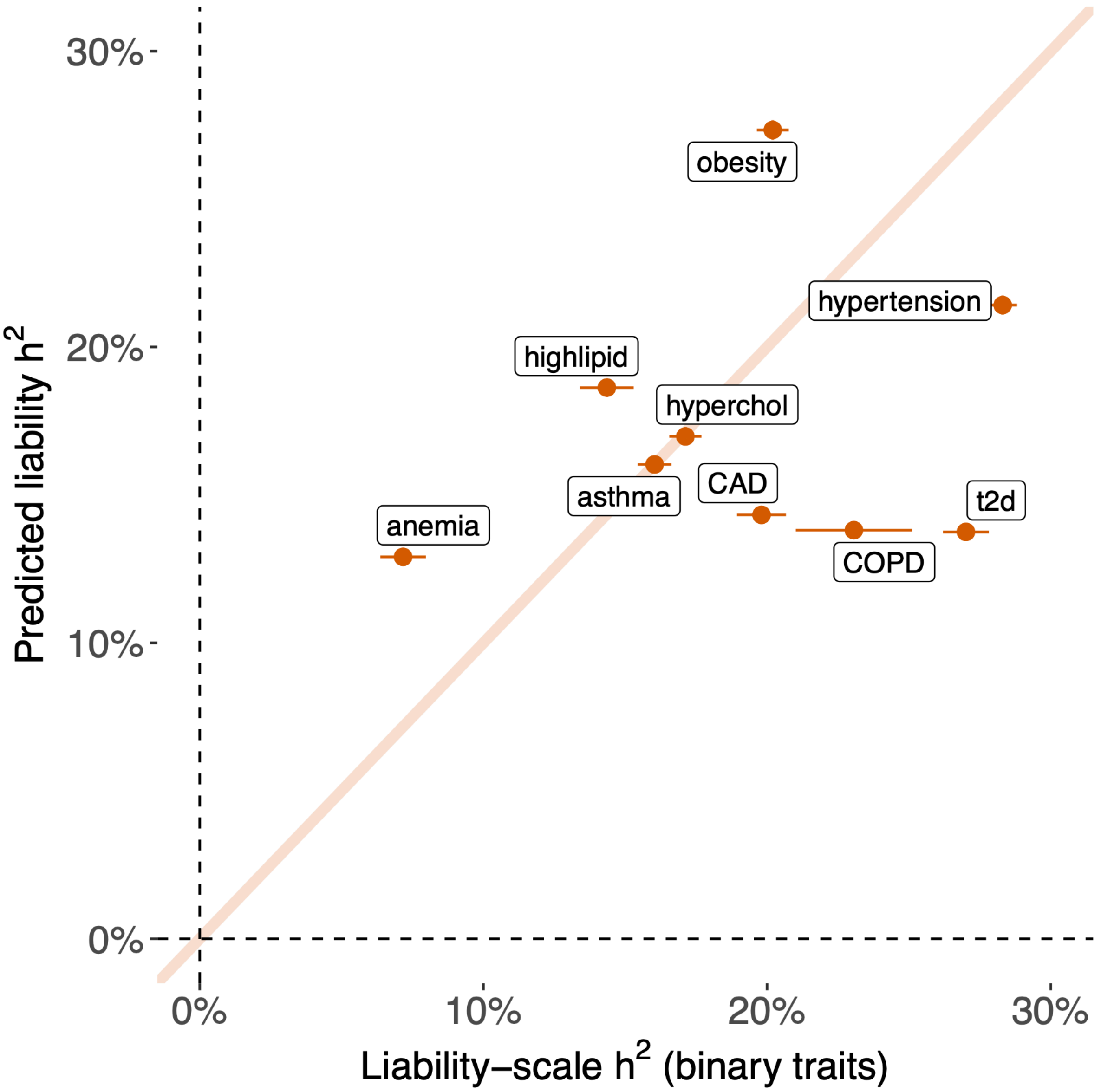


**Supplementary Figure 7. Comparison of heritability of predicted liability and liability-scale heritability of the respective diseases.** Heritability was estimated using BOLT-REML. For diseases, observed-scale heritability is transformed to liability-scale following ref. 14. Labels are the disease name and bars show standard error.

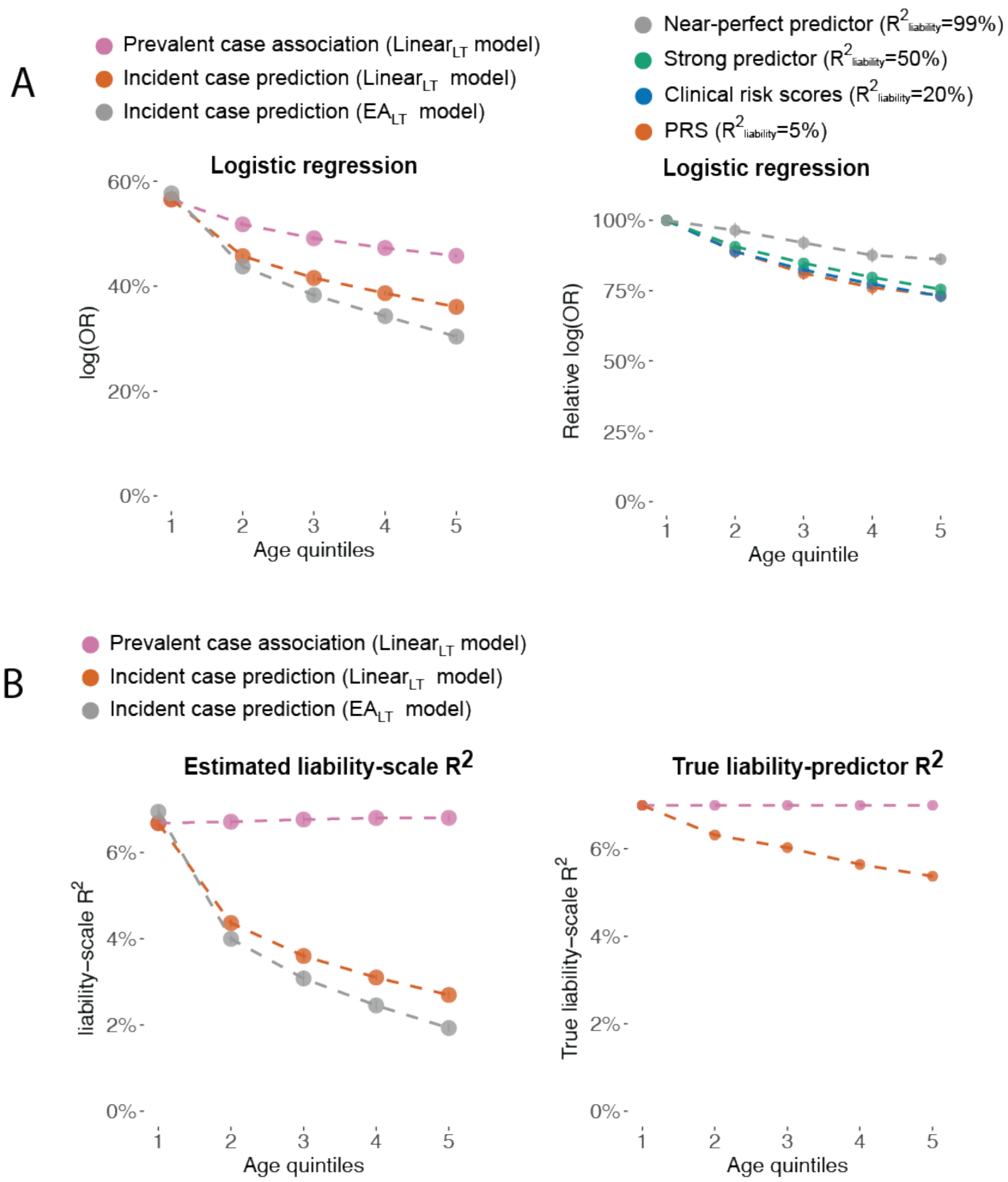


**Supplementary Figure 8. Additional simulations of liability threshold models.** (A) Left: Prevalent case association accuracy (purple), incident case prediction accuracy (red), and Incident case prediction under EA model (grey) across different age quintiles of cases. Prediction accuracy is measured by log odds ratio from logistic regression. Right: Incident case prediction accuracy of predictors with liability-scale $R^2$, measured by log odds ratio from logistic regression. (B) Comparison of estimated liability-scale $R^2$ using Lee et al. 2012 method with the true liability-scale $R^2$ which is estimated by computing square of correlation between the predictor and the simulated liability. For incident case prediction, the correlation is computed within the group that excluded prevalent cases.

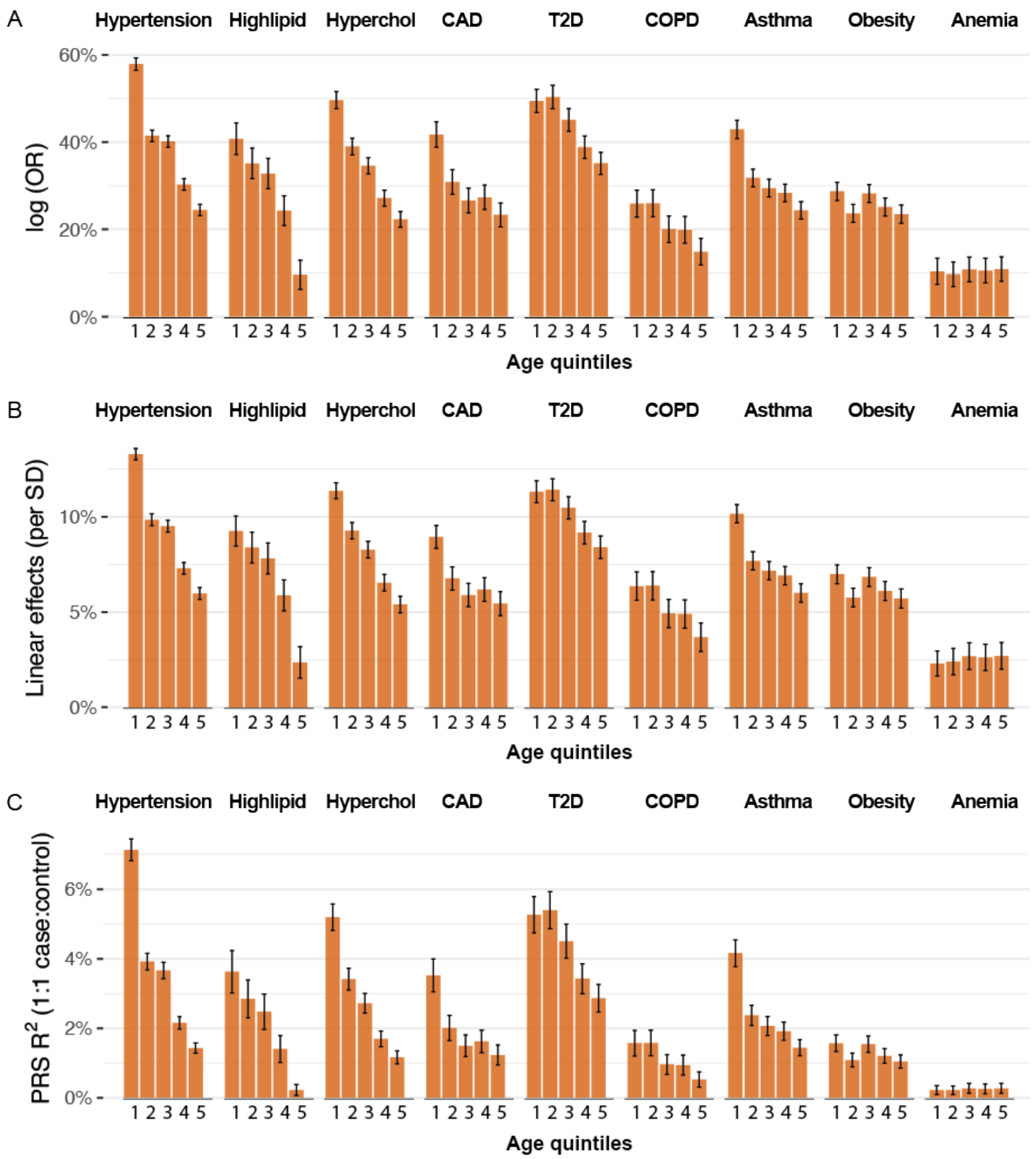


**Supplementary Figure 9. Age-dependent incident case prediction accuracy in 9 diseases.** (A) Log Odds Ratio of PRS on 9 disease traits across 5 age-quintiles. (B) Linear regression coefficient of per-SD PRS on 9 disease traits across 5 age-quintiles. (C) Observed $R^2$ of PRS on 9 disease traits across 5 age-quintiles. Each age bin has the same number of cases and 1:1 case control ratio. Bars show standard error.

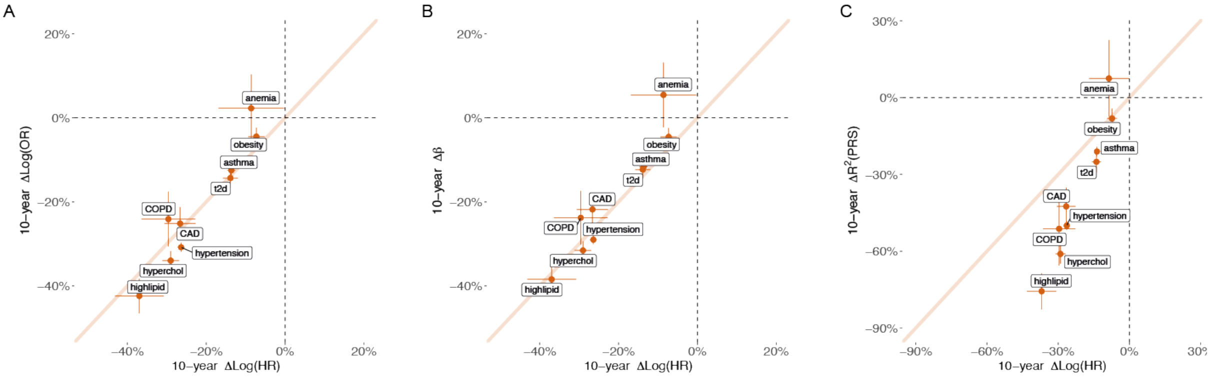


**Supplementary Figure 10. Comparison of four metrics that measures age-dependent incident case prediction accuracy in 9 diseases.** (A) Change of log Odds Ratio of PRS on 9 disease traits w.r.t. to age (per 10 years) compared to change of Log(HR) w.r.t. age. (B) Change of Linear regression coefficient of per-SD PRS on 9 disease traits w.r.t. to age (per 10 years) compared to change of Log(HR) w.r.t. age. (C) Change of Observed-scale $R^2$ of PRS on 9 disease traits w.r.t. to age (per 10 years) compared to change of Log(HR) w.r.t. age. of PRS on 9 disease traits across 5 age-quintiles. Bars show standard error.

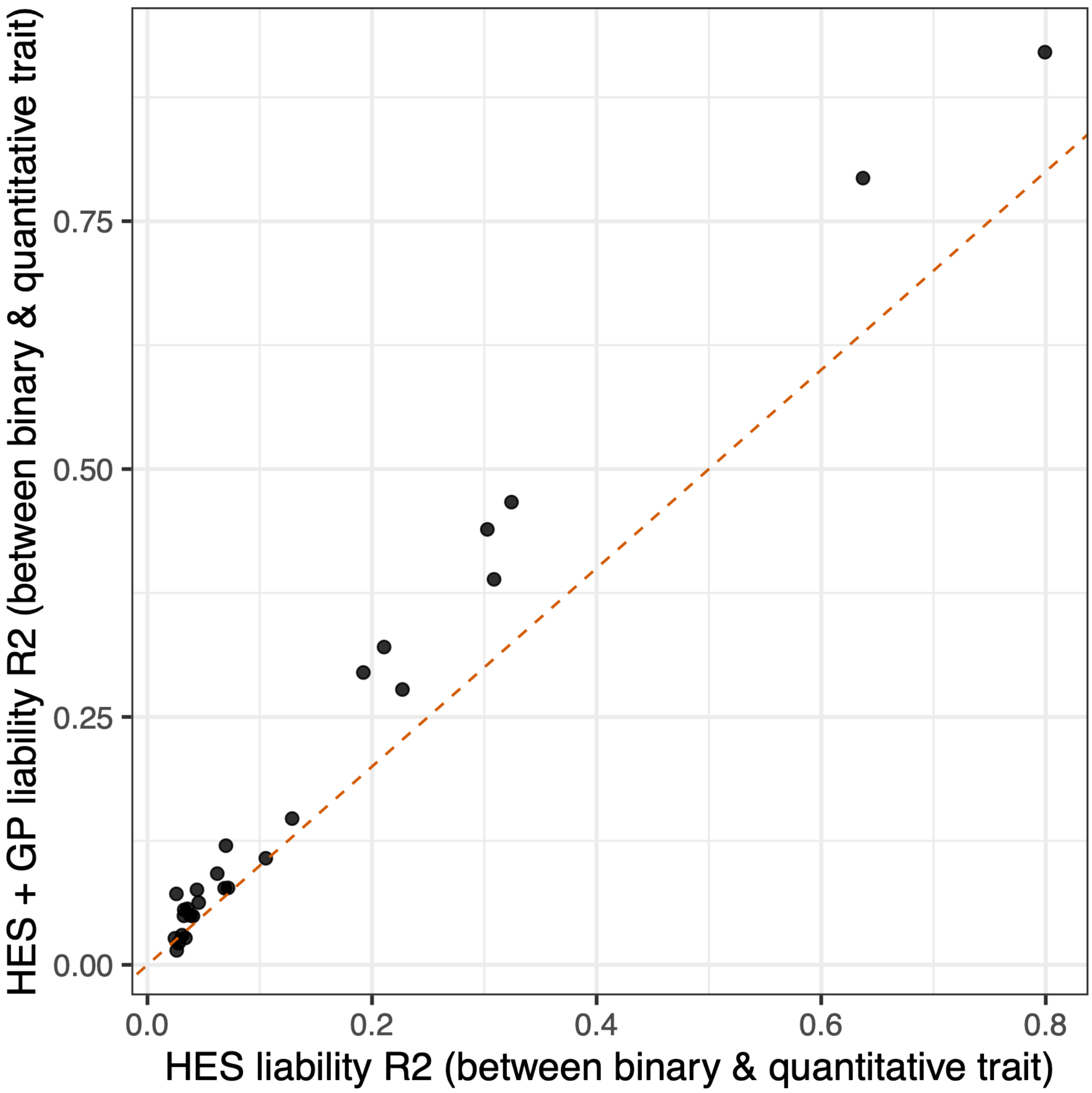


**Supplementary Figure 11. Comparison of using HES and using both HES and GP to define disease cases.** We used two ways to define disease status. First, using diagnoses from hospital inpatient data (HES) to define cases and define others as controls. Second, using the union of diagnoses in HES and primary care data (GP) to define cases and define others as controls. For each disease, we compute its $R_{lib}^2$ with other three quantitative traits that have the strongest correlation with disease status defined using HES only. We then compare the corresponding $R_{lib}^2$ with disease status defined using HES+GP.
